# Impact of natural selection on global patterns of genetic variation, and association with clinical phenotypes, at genes involved in SARS-CoV-2 infection

**DOI:** 10.1101/2021.06.28.21259529

**Authors:** Chao Zhang, Anurag Verma, Yuanqing Feng, Marcelo C. R. Melo, Michael McQuillan, Matthew Hansen, Anastasia Lucas, Joseph Park, Alessia Ranciaro, Simon Thompson, Meghan A. Rubel, Michael C. Campbell, William Beggs, Jibril Hirbo, Sununguko Wata Mpoloka, Gaonyadiwe George Mokone, Regeneron Genetic Center, Thomas Nyambo, Dawit Wolde Meskel, Gurja Belay, Charles Fokunang, Alfred K. Njamnshi, Sabah A. Omar, Scott M. Williams, Daniel Rader, Marylyn D. Ritchie, Cesar de la Fuente Nunez, Giorgio Sirugo, Sarah Tishkoff

## Abstract

We investigated global patterns of genetic variation and signatures of natural selection at host genes relevant to SARS-CoV-2 infection (*ACE2, TMPRSS2, DPP4,* and *LY6E*). We analyzed novel data from 2,012 ethnically diverse Africans and 15,997 individuals of European and African ancestry with electronic health records, and integrated with global data from the 1000GP. At *ACE2,* we identified 41 non-synonymous variants that were rare in most populations, several of which impact protein function. However, three non-synonymous variants were common among Central African hunter-gatherers from Cameroon and are on haplotypes that exhibit signatures of positive selection. We identify strong signatures of selection impacting variation at regulatory regions influencing *ACE2* expression in multiple African populations. At *TMPRSS2*, we identified 13 amino acid changes that are adaptive and specific to the human lineage. Genetic variants that are targets of natural selection are associated with clinical phenotypes common in patients with COVID-19.

## Introduction

Coronavirus disease 2019 (COVID-19) is caused by severe acute respiratory syndrome coronavirus 2 (SARS-CoV-2). Coronaviruses are enveloped, positive-sense, and single-stranded RNA viruses, many of which are zoonotic pathogens that crossed over into humans. Seven coronavirus species, including SARS-CoV-2, have been discovered that, depending on the virus and host physiological condition, may cause mild or lethal respiratory disease. The novel SARS- CoV-2 virus was initially identified in Wuhan, China, in December 2019^1^, and due to high transmission rates, including from asymptomatic subjects^2^, quickly spread globally causing a pandemic of historic proportions. In the US, the crude fatality rate of COVID-19 is ∼ 1%, and mortality increases significantly with age, with 70% of deaths being among individuals 70 years old and above^3; 4^. As is the case with other infectious diseases, COVID-19 progression appears to exhibit sexual-dimorphism, with fatality rates 2-fold greater for men than women^5^. Patients with COVID-19 can be clinically subdivided into three categories: asymptomatic/mild, severe (with dyspnea, hypoxia), and critical (with respiratory failure, shock, or multiorgan dysfunction). The rate of asymptomatic infection of SARS-CoV-2 may be as high as 40-45% ^6^, and those who are asymptomatic are unlikely to convert to acute symptoms even though they may transmit virus for up to 2 weeks. Symptomatic patients may present dry cough, followed by sputum, hyposmia, nasal congestion, nausea, diarrhea, fever and dyspnea, although initial presentation is known to be variable (for example fever or dyspnea may be absent at admission in hospital)^7^. There is considerable variation in disease prevalence and severity across populations and communities. For example, in Chicago, more than 50% of COVID-19 cases and nearly 70% of COVID-19 deaths are in African Americans (who make up 30% of the population of Chicago)^8^. More generally, minority populations in the US appear to have been disproportionally affected by COVID-19; ^8; 9^. In addition, adverse outcomes including death, have been associated with underlying cardiometabolic comorbidities (e.g., hypertension, diabetes, cardiovascular disease, chronic kidney disease)^10–13^. Liver impairment is common in patients with COVID-19, and elevated alanine aminotransferase (ALT) and aspartate aminotransferase (AST) levels are relatively frequent at presentation^2^. The extent to which pre-existing chronic liver conditions affect COVID-19 related complications remains to be elucidated. Smell and taste sensations as well as increased incidence of ischemic stroke have been observed in individuals with COVID-19^14–17^.

Several host genes play a role in SARS-CoV-2 infection^18^. The *ACE2* gene, encoding the angiotensin-converting enzyme-2 protein, was reported to be a main binding site for SARS-CoV during an outbreak in 2003, and evidence showed stronger binding affinity to SARS-CoV-2, which enters the target cells via ACE2 receptors^18; 19^. The *ACE2* gene is located on the X chromosome, its expression level varies among populations^20^, and it is ubiquitously expressed in the lung, blood vessels, gut, kidney, testis, and brain, all organs that appear to be affected as part of the COVID-19 clinical spectrum. SARS-CoV-2 infects cells through a membrane fusion mechanism, which, in the case of SARS-CoV, is known to induce down-regulation of *ACE2*^21^. Such down-regulation has been shown to cause inefficient counteraction of angiotensin II effects, leading to enhanced pulmonary inflammation and intravascular coagulation^21^. Additionally, altered expression of *ACE2* has been associated with cardiovascular and cerebrovascular disease, which is highly relevant to COVID-19 as several cardiovascular conditions are associated with severe disease. Type II transmembrane serine protease (*TMPRSS2*), located on the outer membrane of host target cells, binds to and cleaves ACE2, resulting in activation of spike proteins on the viral envelope, and facilitating membrane fusion and endocytosis^22^. Two additional genes, dipeptidyl peptidase (*DPP4*), and lymphocyte antigen 6 complex locus E (*LY6E*), have been shown to play an important role in the entry of SARS-CoV2 virus into host cells. *DPP4* is a known functional receptor for the Middle East Respiratory Syndrome coronavirus (MERS-CoV), causing a severe respiratory illness with high mortality^23^. Lastly, *LY6E* (lymphocyte antigen 6 complex, locus E) encodes a glycosylphosphatidylinositol (GPI)- anchored cell surface protein which is a critical antiviral immune effector that controls coronavirus infection and pathogenesis^24^. Mice lacking *LY6E* in hematopoietic cells were susceptible to murine coronavirus infection^24^.

In this study, we characterized genetic variation at *ACE2*, *TMPRSS2*, *DPP4*, and *LY6E* in ethnically diverse human populations by analyzing 2,012 novel genomes from ethnically diverse Africans (referred to as the “African Diversity” dataset), 2,504 genomes from the 1000 Genomes project, and whole exome sequencing of 15,997 individuals of European and African ancestry from the Penn Medicine BioBank (PMBB) dataset. The African diversity dataset includes populations with diverse subsistence patterns (hunter-gatherers, pastoralists, agriculturalists) and speaking languages belonging to the four major language families in Africa (Khoesan, Niger-Congo (of which Bantu is the largest subfamily), Afroasiatic, and Nilo-Saharan). We identify functionally relevant variation, compare the patterns of variation across global populations, and provide insight into the evolutionary forces underlying these patterns of genetic variation. In addition, we perform an association study using the variants identified from whole-exome sequencing at the four genes (*ACE2*, *TMPRSS2*, *DPP4*, and *LY6E*) and clinical traits derived from electronic health record (EHR) data linked to the subjects enrolled in the Penn Medicine BioBank (PMBB). The EHR data includes diseases related to organ dysfunctions associated with severe COVID-19 such as respiratory, cardiovascular, liver and renal complications. Our study of genetic variation in SARS-CoV-2 receptors and their partners provides novel data to investigate infection susceptibility within and between populations and indicates that variants in these genes may play a role in comorbidities relevant to COVID-19 severity.

## Results

### Coding variation at *ACE2* among global populations

SARS-CoV-2 employs *ACE2* as a receptor for cellular entry^18^. To systematically characterize genetic variation in the coding region of *ACE2* across global populations, we analyzed whole-genome sequence data from 2,012 individuals from diverse African ethnic groups (referred to as “African diversity panel (ADP)”), 2,504 samples from the 1KG project ^25^, and whole exome sequence data from 15,977 individuals of European and African ancestry from the Penn Medicine Biobank (PMBB) (Figure S1 and Table S1). In total, we identified 41 amino acid changing variants (Figure 1A, and Table S2-3). Twenty-eight (69%), twenty (49%), eighteen (44%), and sixteen (40%) of the nonsynonymous variants were predicted to be deleterious or likely deleterious by the CADD^26^, SIFT ^27^, PolyPhen ^28^ and Condel prediction ^29^ methods (Table S3).

**Figure 1.**
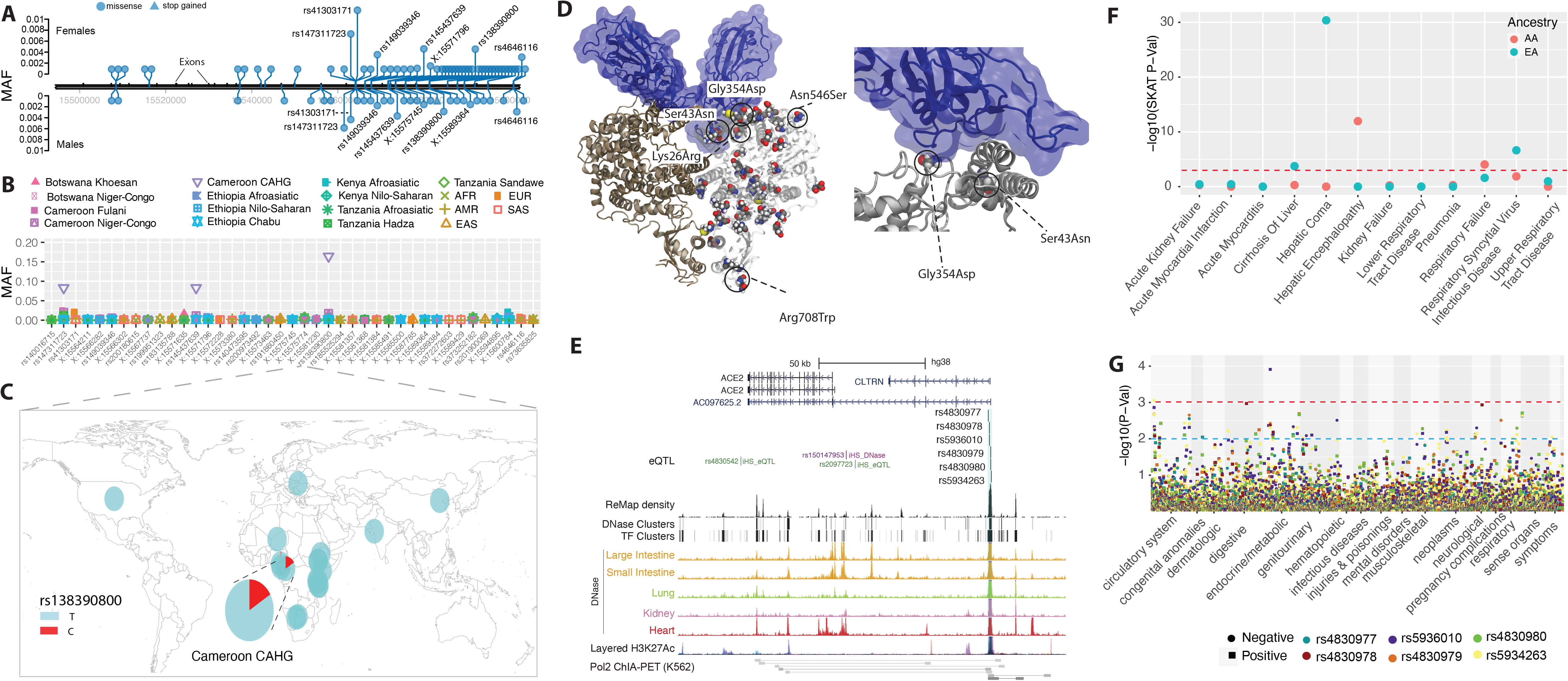
Genetic variation at *ACE2* and its disease association. (A) Location of coding variants and their minor allele frequency (MAF) at *ACE2* identified from the pooled dataset. (B) MAF of coding variants in diverse global ethnic groups. (C) The geographic distribution of the MAF for variants within rs138390800 at *ACE2* in diverse global ethnic groups is highlighted. Each pie denotes frequencies of alleles in the corresponding population. (D) Locations of identified non-synonymous variants within the secondary structure of the *ACE2* protein. (E) Six regulatory eQTLs located in an upstream enhancer of *ACE2*. RNA Pol2 ChIA-PET data and DNase-seq data of large intestine, small intestine, lung, kidney and heart are from ENCODE ^81^. (F) Gene-based association result between coding variants at *ACE2* and 12 disease classes. The disease severity is shown on the x-axis and the y-axis represents the p-values. EA, European Ancestry; AA, African American ancestry. (G) PheWAS plot of six eQTL associated with *ACE2* and ∼1800 disease codes across 17 disease categories. The disease categories are shown on the x-axis and the y-axis represents the -log10 of the p- values. The colored dot represents an eQTL and the direction of effect of the association. The red dashed line denotes the 0.0001 cutoff, and the blue dashed line represent the 0.001 cutoff.

Among the 41 coding variants identified at *ACE2*, the majority are rare (minor allele frequency, MAF < 0.05) in the pooled global population dataset (Figure 1A and Table S3). However, there are variants that are common (MAFs >= 0.05) in the Central African Hunter Gatherer (CAHG) population from Cameroon (often referred to as “pygmies”) (Figure 1B). One of these variants, rs138390800 (Lys341Arg), is a deleterious non-synonymous variant, and present at high frequency (MAF = 0.164) in the CAHG, while it is rare in other African populations and absent in non-African populations (Figure 1C). Two other nonsynonymous variants, rs147311723 (Leu731Phe) (MAF = 0.083) and rs145437639 (Asp597Glu) (MAF = 0.083), are also common only in the CAHG population (Figures 1B and Table S3). These three non-synonymous variants are the only common coding variants found at *ACE2* in any of the populations examined.

We then investigated the potential role of these 41 coding variants in the conformation of the *ACE2* protein. The 41 coding variants are distributed across the entire *ACE2* protein (Figure 1D and Table S3), including its receptor-binding domain (RBD) region which binds to the SARS-CoV-2 spike protein, dimerization interface, and transmembrane helix. In particular, two novel non-synonymous variants Gly354Asp (chrX:15581230) and Ser43Asn (chrX:15600784) are both found directly in the RBD binding region of *ACE2* (Figure 1D and Table S3); the former is only found in low frequency in one population, the Fulani from Cameroon (MAF = 0.008), and the latter is also an African specific variant that is at low frequency in only three East African populations, two of which are Afroasiatic speaking populations from Kenya (MAF = 0.031) and Ethiopia (MAF = 0.012) (Table S3). The variant Arg708Trp (rs776995986) occurs in the region identified as the *TMPRSS2* cleavage site in *ACE2*^30^ and is found only in the Afroasiatic speaking populations from Ethiopia (MAF = 0.004). Importantly, the presence of Arginine residues has been shown to be important in “multibasic” cleavage sites^18^. Therefore, due to the drastic change in physicochemical properties of the residue, this variation could be expected to interfere in *TMPRSS2* cleavage efficiency, though it warrants experimental validation. Finally, two variants are located at glycosylation sites. Variant Asn546Ser (rs756905974, chrX:15572228), which causes the loss of a conserved glycosylation site on the *ACE2* protein, is found only in the SAS populations (MAF = 0.001). Variant Lys26Arg (rs4646116), found in individuals from the European (EUR) (MAF = 0.005), African (AFR) (MAF = 0.001) and South Asian (SAS) (MAF = 0.002) populations from the 1KG dataset (Table S3), occurs near both the conserved *ACE2* glycosylation site Asn90 and the RBD binding site. The modification to a similarly positively charged positive residue could suggest a role for electrostatic interactions, though no direct interference with RBD binding could be deduced without further studies.

### Regulatory variation at *ACE2* among global populations

In contrast to coding variants which have direct effects on protein structure in all cells expressing a gene, the effects of regulatory genetic variants are relatively difficult to determine^31^. Expression quantitative trait locus (eQTL) analysis has been used to identify genetic variants associated with gene expression. We first extracted 2,053 eQTLs significantly associated with *ACE2* gene expression (P < 0.001) from the GTEx database ^32^ (Table S4). To narrow down candidate functional variants, we focus on the eQTLs located in the promoter regions of target genes or in enhancers supported by chromatin interaction data^33^.

We identified six eQTLs (rs4830977, rs4830978, rs5936010, rs4830979, rs4830980 and rs5934263) located in a strong DNase peak at 73.3 kb upstream of *ACE2* that have direct interactions with *ACE2* based on RNA Pol2 ChIA-PET data (Figure 1E, S2 and Table S4). All six SNPs are eQTLs of *ACE2* and all of them have positive normalized effect sizes (NES > 0.2) and significant p-values (P < 0.00008) in brain, tibial nerve, tibial artery, pituitary and prostate cells (Figure S3 and Table S4). In non-African populations, these six eQTLs are in high LD (R^2^ = 0.91 – 1.0) (Figure S4) and, thus there are two common haplotypes: “CCGGAT” and “ATCATC”. The frequency for the “ATCATC” haplotype ranges from 0.31 - 0.47 in all populations except the East Asian population, which has a frequency of 0.068 at all 6 SNPs (Figure S5). In African populations, LD is lower (R^2^ > 0.5; Figure S4), and there are three common haplotypes: “CCGGAT” (0.564), “ATCATC” (0.308), and “CCCGAC” (0.116). Of note, every allele in the haplotype “CCGGAT” is correlated with higher expression of *ACE2* in the cortex of the brain while alleles in haplotype “ATCATC” are correlated with lower expression of *ACE2;* other haplotypes have alleles with both positive and negative effect sizes in different tissues (Table S4). Haplotype “CCCGAC” is only present in populations with African ancestry and its frequency is highest in the Botswana Khoesan (0.38) and Cameroon CAHG (0.38) populations. We also identified one variant (rs186029035) located in strong TF and DNase clusters (ENCODE) in the 16^th^ intron of *ACE2*. This variant is only common in the Cameroon CAHG population and, therefore, there is no eQTL data for this SNP in the GTEx database (MAF = 0.153, Table S2).

### Signatures of natural selection at *ACE2*

As indicated above, most of the non-synonymous variants at *ACE2* are rare in global populations and many of them are predicted to be deleterious, indicating that this gene is under strong purifying selection. To formally test for signatures of natural selection at *ACE2*, we first examined the ratio of non-synonymous and synonymous variants at each gene using the dN/dS test^34^. The dN/dS for all pooled samples was 0.77, indicating that *ACE2* is under purifying selection globally (Table S5 and Figure S6). However, in the East Asian population, we observed seven non-synonymous variants (all of them are rare) and only one synonymous variant, and the dN/dS value is 1.85, indicating an excess of non-synonymous variation. In other populations, the dN/dS ratio ranges from 0 to 0.79 (Table S5 and Figure S6). Thus, *ACE2* appears to be under strong purifying selection in most populations but may be under weak purifying selection in the East Asian population. We next applied the MK-test^35^ which compares the ratio of fixed non-synonymous sites between humans and chimpanzee (Dn = 8) and fixed synonymous sites (Ds = 6) to the ratio of polymorphic nonsynonymous sites among populations (Pn = 41) relative to polymorphic synonymous sites (Ps =14) and found that it is not significant (odds ratio (OR) = 0.45, P = 0.94, two-sided Fisher’s exact test, Table S6 and Figure S7, S8).

Because the above-mentioned methods are more suitable for detecting signals of natural selection acting over long time scales^36; 37^, we then tested for signatures of recent positive selection at *ACE2* in global populations using the iHS test^38^ to detect extended haplotype homozygosity (EHH)^39^, which identifies regions of extended linkage disequilibrium (LD) surrounding a positively selected locus. We first focused on the three common non-synonymous variants in the CAHG population from Cameroon (rs138390800, rs147311723 and rs145437639; MAF: 0.083 - .164), and a common putative regulatory variant (rs186029035, located in TF and DNase clusters in the 16^th^ intron of *ACE2,* MAF = 0.153). The derived alleles of these variants exist on three different haplotype backgrounds: rs147311723 and rs145437639 are on the same haplotype backgrounds, while rs138390800 and rs186029035 are on similar, but distinct, haplotype background (Figure 2A). The derived alleles of the corresponding SNPs on each haplotype background show EHH extending longer than 2 Mb, while the ancestral alleles of these SNPs harbor haplotypes extending less than 0.3 Mb (Figure 2B). We then calculated the integrated haplotype score (iHS) of each of these variants, to determine whether these extended haplotypes are unusually long compared to other SNPs with a similar allele frequency; the iHS values were not significant for any of these variants (Figure S9 and Table S7). However, if selection were acting on multiple haplotypes simultaneously (as shown above), the EHH and iHS tests would not be well powered to detect selection^40^. We also used the *d_i_* statistic^41^ to measure if allele frequencies at these candidate SNPs were strongly differentiated between Cameroon CAHG and other populations. The *d_i_* values of SNPs rs138390800 and rs186029035 were in the top 1.4% and 1.7%, respectively, of *d_i_* values for all SNPs examined, indicating that that allele frequencies at these variants are amongst the most highly differentiated in the CAHG population. However, it should be noted that in the CAHG these four variants (rs138390800, rs147311723, rs145437639 and rs186029035) are in complete LD based on D’ (D’ = 1) with the 6 eQTLs described above (Figure S10, S11), indicating that the alleles are on the same haplotype background. Thus, it is not possible to distinguish if the non-synonymous variants are targets of selection or if they are “hitchhiking” to high frequency due to selection on flanking regulatory variants. Given the high LD in the region, it is possible that multiple functional variants on the same haplotype backgrounds have been under selection.

**Figure 2.**
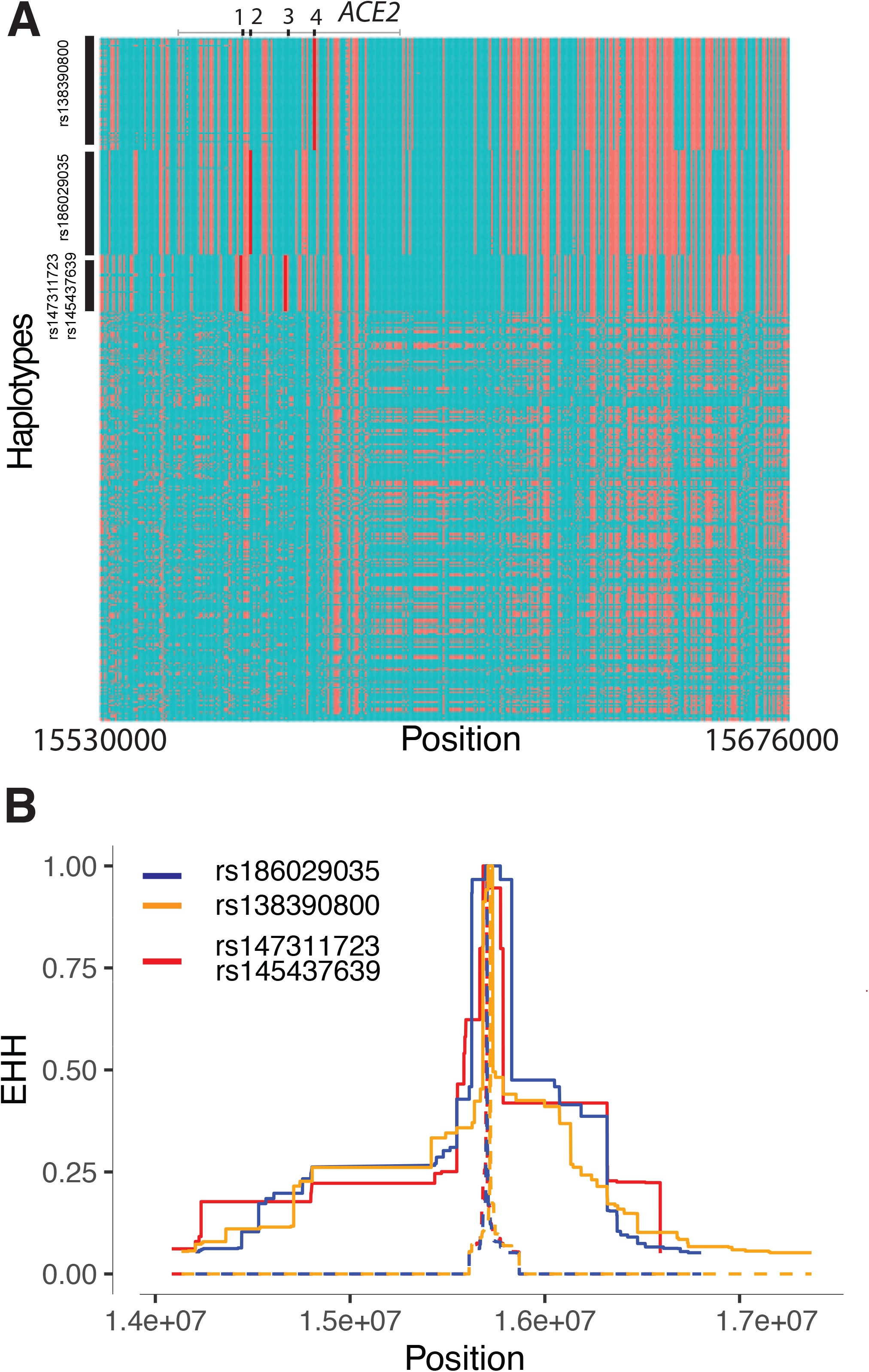
Natural selection signatures at *ACE2* in the Cameroon CAHG populations. (A) Haplotypes over 150kb flanking *ACE2* in CAHG populations. The X-axis denotes genetic variant position, and the y-axis represents haplotypes. Each haplotype (one horizontal line) is composed of the genetic variants (columns). Red dots indicate the derived allele, while green dots indicate the ancestral allele. Haplotypes surrounded by a top-left vertical black line suggest these haplotypes carry derived allele(s) of the labeled variant near the corresponding black line. For example, the first black line denotes all the haplotypes that have the derived allele at rs138390800 (dark red line). Haplotypes carrying rs138390800, rs147311723, rs145437639, and rs186029035 show more homozygosity than other haplotypes. 1, 2, 3, 4 at the top of the plot denotes positions for rs147311723, rs186029035, rs145437639 and rs138390800, respectively. (B) Extended haplotype homozygosity (EHH) of rs138390800, rs186029035 and rs147311723 (rs145437639 is in strong LD with rs147311723) at *ACE2* in CAHG populations.

We then investigated signatures of recent positive selection at candidate regulatory variants near *ACE2* in the global datasets. In total, there are 234 variants that had high iHS scores (|iHS| > 2) in at least one population extending over an ∼200 kb region (Table S8), and 48% (n=113) of these variants are either eQTLs or located at DNase hypersensitive regions which are in high LD based on D’ (Figure S10, S11, and Table S8). Among the region near the TSS (<10kb from *ACE2*), there are two variants in high LD (D’ = 1) that had high iHS scores in the San population from Botswana (Figure 3A); rs150147953 is located in a DNase peak in multiple tissues including lung, intestine and heart and rs2097723 is an eQTL of *ACE2* in the brain (Figure 1E, S12; Table S8). We also identified strong selection signals at the region 50 – 120 kb upstream of *ACE2* (chrX: 15650000-15720000) in the AFR, San from Botswana, and Niger-Congo-speaking populations from Cameroon, as well as Afroasiatic- and Nilo-Saharan-speaking populations from Kenya (Figure 3A and S8). Two SNPs in this region (rs5936010 and rs5934263) have elevated iHS scores (|iHS| > 2) in the San population from Botswana and the Afroasiatic population from Kenya (Figure 3A) and are part of the 6 eQTLs described above, located within a strong enhancer interacting with the promoter of *ACE2* (Figure 1E). Two additional eQTLs, that are in complete LD with the 6 eQTLs (D’ = 1; Figure S11), rs4830984 and rs4830986, had high iHS scores in four of the five African populations listed above (all but the Kenya Afroasiatic; Figure 3A).

**Figure 3.**
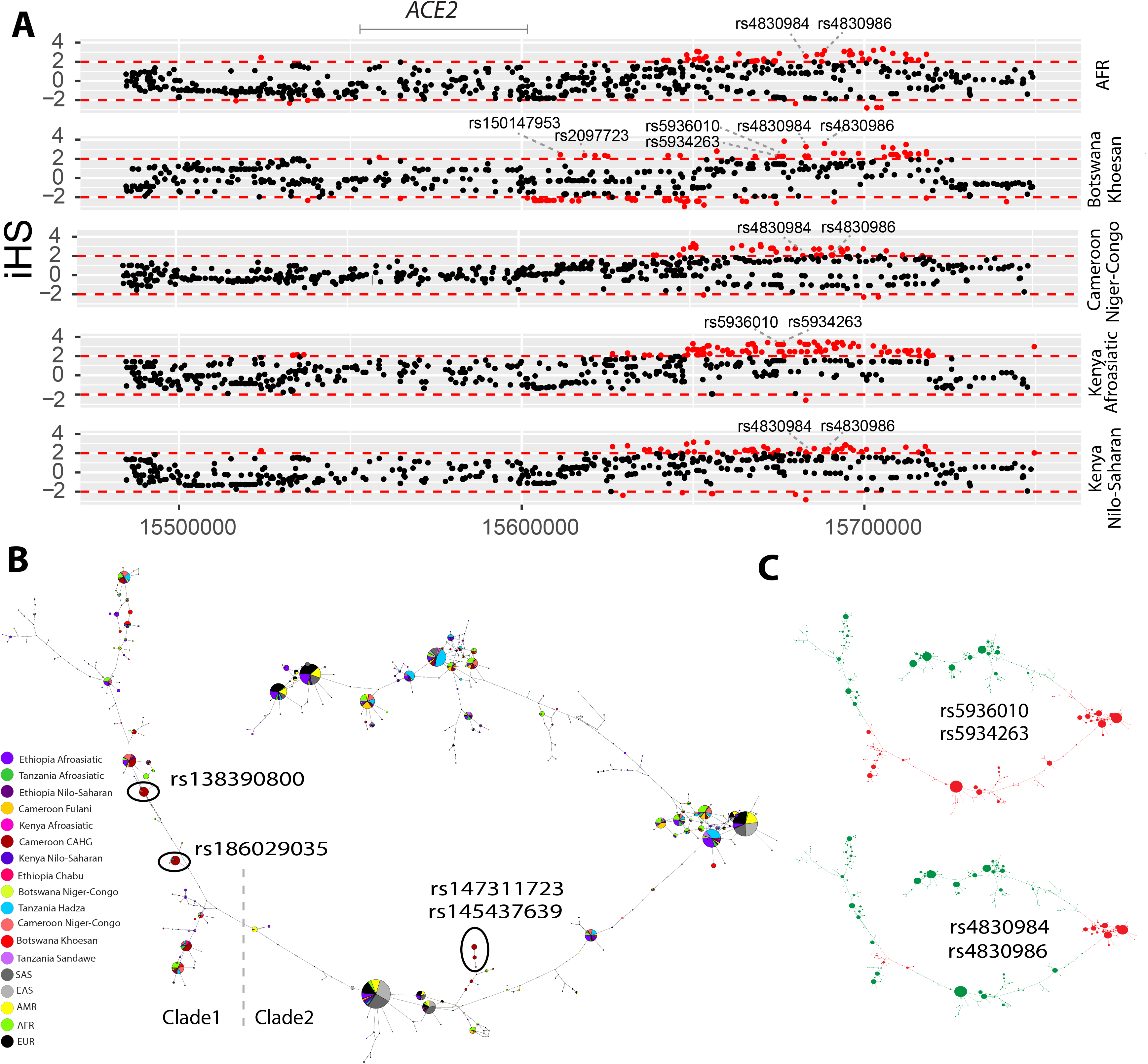
Natural selection signatures at the upstream region of *ACE2* in African populations. (A) iHS signals at the upstream region of *ACE2* (chrX:15650000-15720000) in African populations. Each dot represents a SNP. Red dots denote SNPs that are significant (|iHS|>2). The gray solid line denotes the gene body region of *ACE2*. Putatively causal tag SNPs were annotated in the plots. (B) Haplotype network over 150kb flanking *ACE2* in diverse ethnic populations. The network was constructed with SNPs that showed iHS signals in all populations and overlapped with DNase regions or eQTLs. The four functional candidates identified in Cameroon CAHG were also included in the networks. Each pie represents a haplotype, each color represents a geographical population, and the size of the pie is proportional to that haplotype frequency. In the left panel, dashed line denotes the boundary of clade 1 and clade 2. Black oval denotes haplotypes containing the corresponding variants. (C) Haplotype containing variants (rs5936010, rs5934263, rs4830984 and rs4830986) are highlighted. Red pie denotes haplotypes containing the derived allele of the corresponding variants, while green pie denotes haplotypes containing the ancestral allele of the corresponding variants

We performed haplotype network analysis to examine phylogenetic relationships among haplotypes at *ACE2* in global populations derived for SNPs showing signatures of natural selection (Figure 3B and 3C). We identified two haplotype clades: one (clade 1) is nearly specific to Africans and the other (clade 2) encompasses global populations (Figure 3B). In the CAHG, haplotypes containing the rs138390800 (Lys341Arg) non-synonymous variant and the rs186029035 regulatory variant are in clade 1, whereas haplotypes containing the rs147311723 (Leu731Phe) and rs145437639 (Asp597Glu) non-synonymous variants are located in clade 2 (Figure 3B). Haplotypes containing the two regulatory variants (rs5936010 and rs5934263) located 50 – 120 kb upstream of *ACE2* are shared in global populations, and the nearby regulatory variants rs4830984 and rs4830986 are sub-lineages on those haplotype backgrounds (Figure 3B and 3C).

### Associations between genetic variations in *ACE2* and clinical disease phenotypes

We examined associations of genetic variation at *ACE2* with clinical phenotypes using the PMBB cohort that consists of exome-sequencing data from 15,977 participants between the ages of 19 and 89 years (52% female) with extensive clinical data available through their electronic health records (EHR). Of these, 7061 individuals were of European ancestry (42%) and 8916 were of African ancestry (55%) (Table S1).

To test for association between rare coding variants and clinical phenotypes, we applied a gene-based approach^42; 43^ and single variant analysis. First, we performed a gene-based analysis by collapsing the coding region variants with MAF < 0.01 that are annotated as non-synonymous or putative loss-of-function (pLOF) variants. We tested for association with 12 phenotypes, encompassing COVID-relevant disease classes affecting different organ systems, defined by EHR based diagnosis codes (Table S9). For the gene-based approach, we applied two statistical tests: a) a burden test (i.e. the cumulative effect of rare variants in a gene) that uses logistic regression and b) a sequence kernel association test (SKAT)^44^. Thus, it can compute effect estimates but may suffer from loss of power when gene variants have effects in opposite directions (i.e., protective and higher risk variants). This limitation can be overcome by parallel analysis with SKAT, a powerful approach to model mixed effect variants. However, this approach does not provide effect estimates. Therefore, we reported outcomes using both methods. Ancestry specific analysis of gene-based tests identified seven associations in African ancestry (AA) and three associations in European ancestry (EA) populations that reached statistical significance levels after multiple hypothesis correction (p < 1 x 10^-04^) for the SKAT model. None of the gene burden models reached a significance level of p < 1 x 10^-04^. The effect size from the logistic regression model was used to indicate a protective or increased risk effect on disease phenotype. In the AA population, the most significant associations were with hepatic encephalopathy and respiratory failure (Figure 1F and Table 1). The association with respiratory failure is interesting as it is one of the key severe clinical features reported for COVID-19^10; 45–48^. However, the same association was not significant in the EA population, which could be explained by lack of power due to lower number of coding variants at *ACE2* in EA. Within the EA population, the most significant associations included hepatic coma, respiratory syncytial virus infectious disease, and cirrhosis of the liver (Table 1).

**Table 1.** Associations of ACE2, DPP4, TMPRSS2, and LY6E with 12 disease classes derived from EHR data.

| Disease Phenotype | Gene | Cases | Controls | Carrier Controls | Carrier Cases | SKAT P | Burden P | Burden OR | Burden SE | 95% CI | Dataset |
| --- | --- | --- | --- | --- | --- | --- | --- | --- | --- | --- | --- |
| Hepatic Encephalopathy | *ACE2* | 97 | 8045 | 441 | 5 | 1.1E-12 | 0.0043 | 5.73 | 0.61 | 0.55 - 2.94 | AA |
| Respiratory Syncytial Virus Infectious Disease | *DPP4* | 56 | 6392 | 85 | 1 | 6.8E-07 | 0.1221 | 6.06 | 1.17 | -0.48 - 4.09 | AA |
| Respiratory Failure | *TMPRSS2* | 199 | 6392 | 11 | 2 | 2.3E-06 | 0.0124 | 7.31 | 0.80 | 0.43 - 3.55 | AA |
| Respiratory Failure | *ACE2* | 199 | 6392 | 351 | 12 | 9.0E-05 | 0.0509 | 3.10 | 0.58 | 0 - 2.26 | AA |
| Upper Respiratory Tract Disease | *DPP4* | 144 | 6392 | 85 | 3 | 2.5E-04 | 0.0978 | 4.16 | 0.86 | -0.26 - 3.11 | AA |
| Respiratory Syncytial Virus Infectious Disease | *TMPRSS2* | 56 | 6392 | 11 | 1 | 3.9E-04 | 0.0217 | 11.63 | 1.07 | 0.36 - 4.55 | AA |
| Pneumonia | *LY6E* | 1120 | 6392 | 7 | 5 | 1.0E-02 | 0.0108 | 6.09 | 0.71 | 0.42 - 3.19 | AA |
| Respiratory Syncytial Virus Infectious Disease | *ACE2* | 56 | 6392 | 351 | 7 | 1.3E-02 | 0.1857 | 3.85 | 1.02 | -0.65 - 3.34 | AA |
| Lower Respiratory Tract Disease | *TMPRSS2* | 693 | 6392 | 7 | 1 | 3.4E-02 | 0.1541 | 2.59 | 0.67 | -0.36 - 2.26 | AA |
| Pneumonia | *TMPRSS2* | 1120 | 6392 | 11 | 4 | 4.8E-02 | 0.2029 | 2.23 | 0.63 | -0.43 - 2.04 | AA |
| Hepatic Coma | *ACE2* | 16 | 6817 | 318 | 1 | 4.3E-31 | 0.0019 | 10.45 | 0.76 | 0.87 - 3.83 | EA |
| Respiratory Syncytial Virus Infectious Disease | *ACE2* | 40 | 5859 | 274 | 3 | 2.3E-07 | 0.1650 | 3.61 | 0.92 | -0.53 - 3.1 | EA |
| Cirrhosis Of Liver | *ACE2* | 10 | 6817 | 43 | 1 | 1.8E-04 | 0.0837 | 9.40 | 1.30 | -0.3 - 4.78 | EA |
| Acute Myocardial Infarction | *LY6E* | 396 | 6494 | 8 | 1 | 1.8E-02 | 0.2936 | 3.65 | 1.23 | -1.12 - 3.71 | EA |

We also examined an extended list of ∼1800 phecodes derived from the EHR and 33 EHR-based quantitative lab measurements and performed a phenome-wide association study (PheWAS)^42; 43; 49^. After multiple testing correction, we identified one association in the AA and five associations in the EA populations reaching study-wide significance (p < 1 x 10^-5^) (Table S9 and S5). Myocarditis, a rare cardiovascular disease caused by viral infection, was the top PheWAS association in the AA population but not significant in the EA population. Although the population difference for this specific association is unclear, recent studies have reported a link between SARS-CoV-2 induced cardiac injury among COVID-19 patients^50^, which it was suggested might be mediated by *ACE2*. This observation would be consistent with *ACE2* expression in heart tissue, and its upregulation in cardiomyocytes^50–52^. Among respiratory diseases, cough and allergic rhinitis reached nominal significance (p < 0.01) in the AA population (Table S9). In the EA population, we identified a nominal association with influenza, asthma, emphysema, cough, and painful respiration (p-value<0.01). Our findings in the EA cohort are consistent with other studies of *ACE2* in subjects derived from the UK biobank^53^. Among the median measure of 33 EHR-based quantitative lab measurements that we investigated (see Methods), only the internationalized normalized ratio (INR) derived from the prothrombin time test showed a nominal association with increase in INR above 1.11, potentially relevant to blood clotting abnormalities observed in COVID patients (Table 2).

**Table 2.** Association of *ACE2*, *DPP4*, *TMPRSS2*, and *LY6E*with clinical laboratory measures derived from the EHR.

| Lab Name | Gene | Sample Size | Carriers | Beta | SE | P | Dataset |
| --- | --- | --- | --- | --- | --- | --- | --- |
| Urine Bilirubin | TMPRSS2 | 1410 | 2 | 3.13 | 0.95 | 0.001 | EA |
| Total Cholesterol | LY6E | 5800 | 8 | 41.11 | 15.29 | 0.007 | AA |
| Prothrombin | LY6E | 6220 | 10 | 3.47 | 1.31 | 0.008 | AA |
| Eosinophil (%) | LY6E | 7697 | 12 | -1.34 | 0.56 | 0.016 | AA |
| Eosinophil (THO/uL) | LY6E | 7678 | 12 | -0.09 | 0.04 | 0.022 | AA |
| Prothrombin | LY6E | 5944 | 7 | 5.17 | 2.52 | 0.040 | EA |
| Prothrombin | ACE2 | 6220 | 330 | 1.11 | 0.55 | 0.045 | AA |

To further evaluate the individual effect of each rare coding variant in *ACE2*, we performed a single variant association analysis on the rare variants in the genes identified from gene-based tests. A Fishers exact test was used to account for the small sample size when testing the impact of rare single variants on phenotypes. The *ACE2* variant rs147311723, which is only present in African populations, was most significantly associated with respiratory infection (p <0.05, OR=1.95 [1.06 - 3.6]). Another African specific *ACE2* variant rs138390800 did not reach statistical significance but showed modestly increased risk of respiratory failure (p=0.1; OR=2.29 [0.83 - 6.33]).

For the six eQTLs identified near *ACE2* (rs4830977, rs4830978, rs5936010, rs4830979, rs4830980 and rs5934263), we performed a PheWAS with clinical data by ancestry. We found that two of the six eQTLs (rs5936010 and rs5934263) (targets of positive selection in both Afroasiatic populations from Kenya and Khoesan populations from Botswana) are significantly associated with type 2 diabetes (p=1.23 x 10^-4^, OR=1.1) and hypertension (p=8.8 x 10^-4^, OR=1.13), respectively, in the AA population (Figure 1G, and Table S10). Among the respiratory disorders, all six eQTLs had nominal associations (p < 0.01) with acute sinusitis and dypnea (shortness of breath) in AA and bronchiectasis in EA. Further, we noticed a difference in the effect of association among the six eQTLs with respiratory disorders examined in AA. Variants rs5936010 (OR=1.11) and rs5934263 (OR = 1.19) were associated with increased risk of respiratory disorder, whereas the rest of the four eQTL variants were associated with decreased risk.

### Genetic variation at *TMPRSS2* and its potential role in SARS-COV-s2 infection susceptibility

The trans-membrane protease serine 2 (TMPRSS2) protein enhances the spike protein-driven viral entry of SARS-CoV-2 into cells^18^. At this gene, we identified forty-eight nonsynonymous variants. Among the non-synonymous variants, only two (rs12329760 [Val197Met] and rs75603675 [Gly8Val]) have high MAF (> 0.05) in the pooled global dataset (Figure 4A, and Table S11). While rs75603675 is highly variable in non-East-Asian populations (AFR = 0.3, AMR = 0.27, EUR = 0.4, and SAS = 0.2), it is not highly variable in East Asians (MAF = 0.02) (Figure 4B and 4C, and Table S11). In addition, some non-synonymous variants were common and specific to African populations. Notably, the non-synonymous variant rs61735795 (Pro375Ser) had a high MAF in the Khoesan-speaking population from Botswana (MAF = 0.18). This variant is present at low frequency in populations from Cameroon (MAF < 0.01) and Ethiopia (MAF < 0.03) and was absent in non-African populations. The non-synonymous variant rs367866934 (Leu403Phe) is common in the Cameroonian CAHG population (MAF = 0.15) and has low frequency (MAF = 0.02) in other populations from Cameroon, but it is absent from non-Cameroonian populations (Figure 4B and Table S11). Another non-synonymous variant rs61735790 (His18Arg) is common in the CAHG populations from Cameroon (MAF = 0.12) and the Nilo-Saharan populations from Ethiopia (MAF = 0.12) but is rare in other populations (Figure 4B and Table S11).

**Figure 4.**
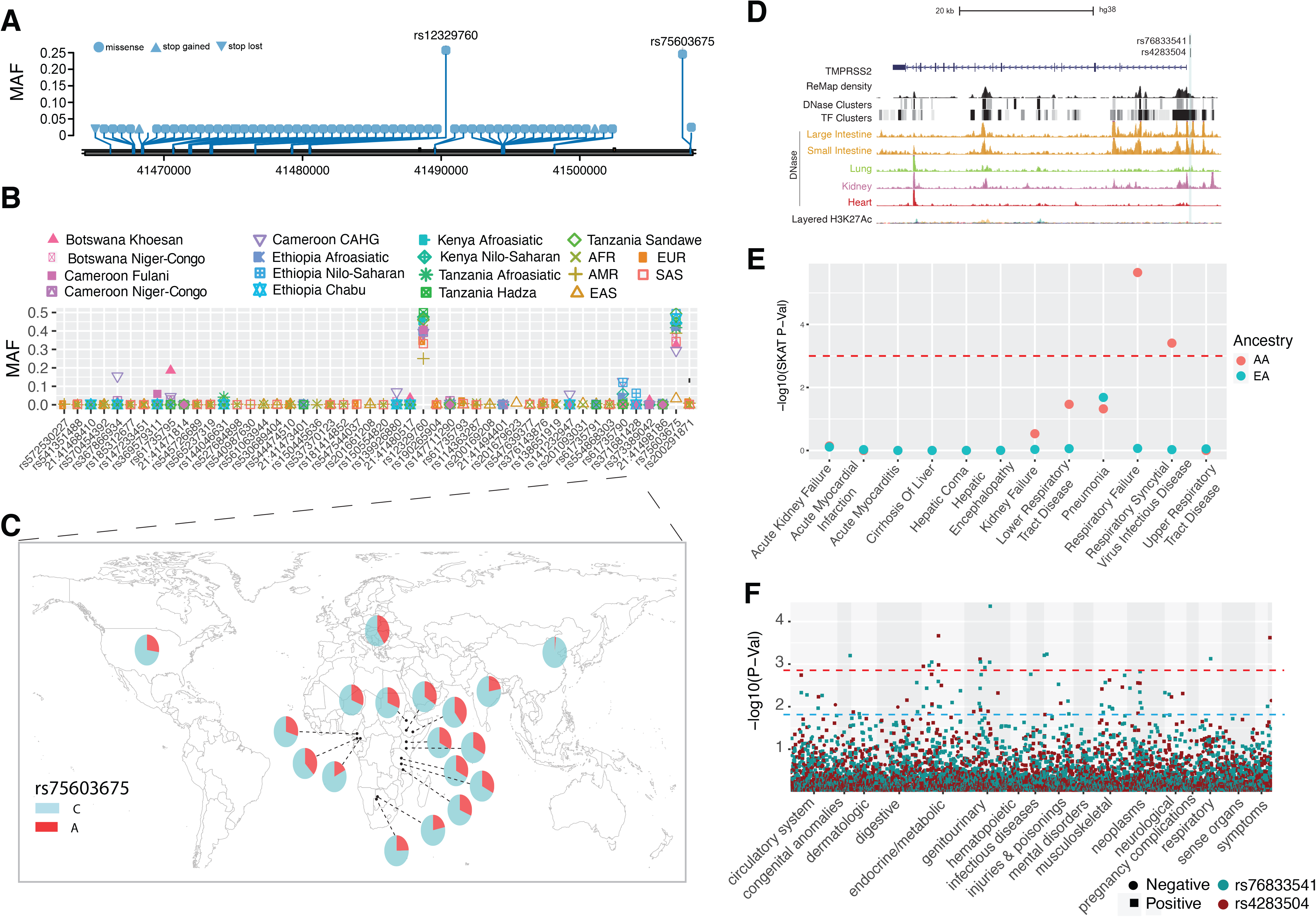
Genetic variation at *TMPRSS2* and its disease association. (A) Location of coding variants and their minor allele frequency (MAF) at *TMPRSS2* identified from the pooled dataset. (B) MAF of coding variants in diverse global ethnic groups. (C) The geographic distribution of MAF of variant within rs75603675 at *TMPRSS2* in diverse global ethnic groups. (D) Two regulatory eQTLs located in the promoter region of the *TMPRSS2* gene. RNA Pol2 ChIA-PET data and DNase-seq data of large intestine, small intestine, lung, kidney and heart are from ENCODE^81^. (E) Gene- based association result between coding variants at *TMPRSS2* and 12 disease classes. The disease classes are shown on the x-axis and the y-axis represents the p-values. EA, European Ancestry; AA, African American ancestry. (F) PheWAS plot of the two eQTLs associated with *TMPRSS2* and ∼1800 disease codes across 17 disease categories. The disease categories are shown on the x-axis and the y-axis represents the -log10 of the p-values. The colored dot represents an eQTL and the direction of effect of the association. The red dashed line denotes the 0.0001 cutoff, and the blue dashed line represents the 0.001 cutoff.

We identified two regulatory SNPs (rs76833541 and rs4283504) in the promoter region of the *TMPRSS2* gene that have been identified as eQTLs of *TMPRSS2* in testis (Figure 4D, S13, and Table S4). The MAF of rs76833541 is higher in EUR (MAF = 0.16) than other populations (EAS = 0.002, AFR = 0.006, AMR = 0.06 and SAS = 0.05) and the MAF of rs4283504 is more common in EAS (MAF = 0.21) than other populations (EUR = 0.11, AFR = 0. 04, AMR = 0.12 and SAS = 0.14) (Figure S14 and Table S2).

### Signatures of natural selection at *TMPRSS2*

We applied the MK test at *TMPRSS2* and observed that Dn/Ds (13/2) is significantly larger than Pn/Ps (48/45) among pooled human samples (OR = 6.1, P-val = 0.009, Fisher’s exact test) (Figure 5A, and Table S6) as well as in individual ethnic groups (OR ranged from 5.0 - 17), indicating positive selection in the hominin lineage after divergence from chimpanzee. Notably, there are 13 non-synonymous and 2 synonymous variants at *TMPRSS2* (ENST00000398585.7, see Figure S15 for ENST00000332149.10) that were fixed in human populations. The non-synonymous variants are located in different structural domains of *TMPRSS2*: amino acid A3P, N10S, T46P, A70V, R103C, and M104T are located in the cytoplasmic region which may function in intracellular signal transduction^54^; L124I is located in the transmembrane region; N144K is located in the extracellular region; S165N and S178G are located in the LDL-receptor class A domain; E441Q and T515M are located in the Peptidase S1 domain which is involved in the interaction with the SARS-CoV-2 spike protein^18^; S529G is located in the last amino acid position of the protein (Figure 5B). In contrast to the MK test, the dN/dS ratio test was not significant in any population, indicating no excess of non-synonymous to synonymous variation within populations. (Table S5 and Figure S6).

**Figure 5.**
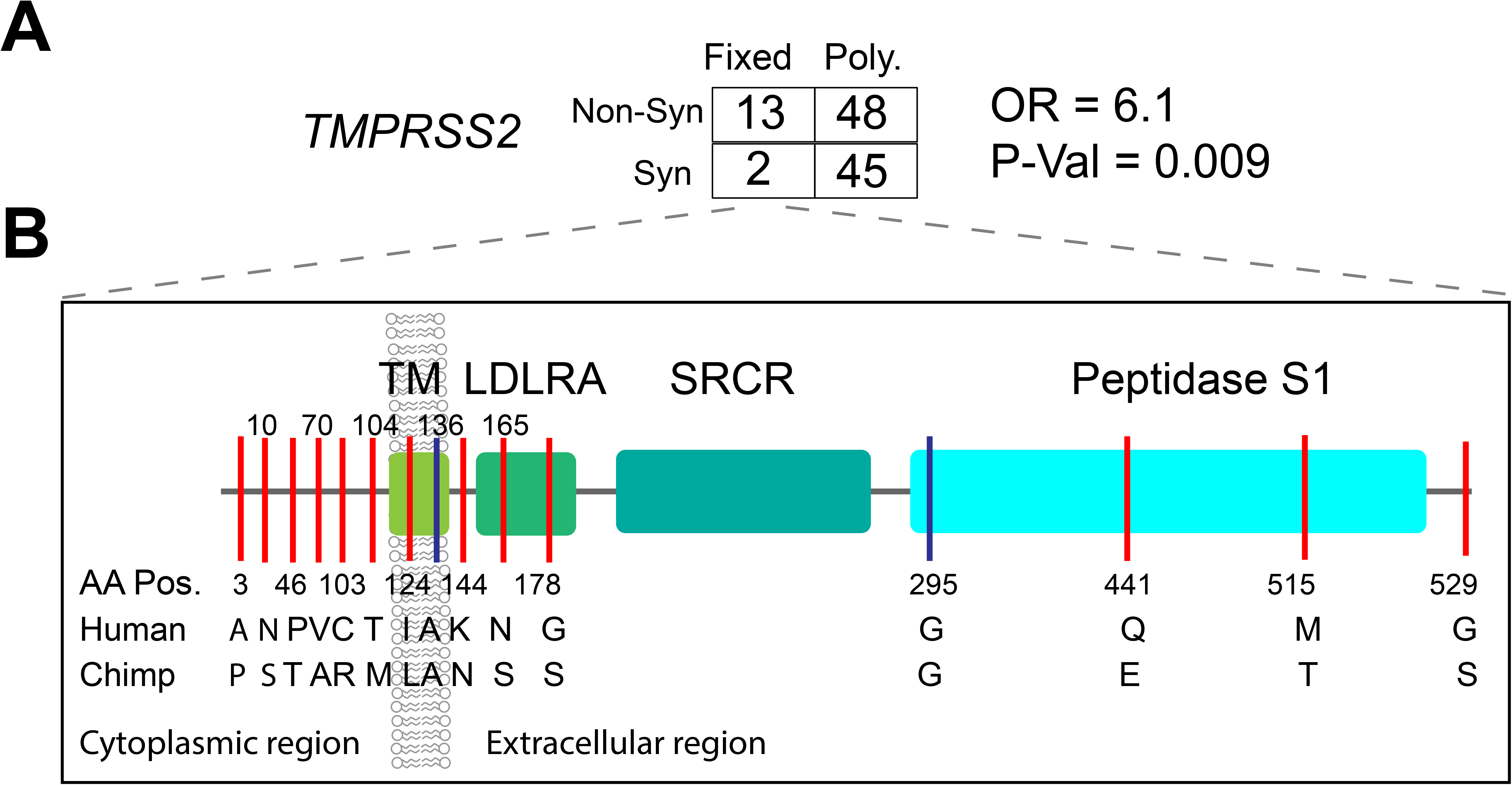
Natural selection signatures of *TMPRSS2*. (A) The result of the MK-test for *TMPRSS2* in the pooled dataset. Non-syn indicates non-synonymous variants; Syn indicates synonymous variants. “Fixed” denotes variants that were fixed between the human and the Chimpanzee; “Poly” represents polymorphic variants within human populations. OR, odds ratio. The transcript *ENST00000398585.7* was used for calculation. (B) Illustration of locations of variants that are divergent between the human and Chimpanzee lineages on the *TMPRSS2* protein domains. Boxes denote the protein domains of *TMPRSS2*. Red lines represent non-synonymous variants that occurred in the corresponding domains of *TMPRSS2*, with the amino acids and positions of the Human and the Chimpanzee annotated at the bottom of the lines. Blue lines denote synonymous variants. TM, transmembrane domain; LDLRA, LDL-receptor class A; SRCR, scavenger receptor cysteine-rich domain 2; Peptidase S1, Serine peptidase.

We also tested for recent positive selection at *TMPRSS2* in all ethnic groups using iHS (Figure S16 and Table S7). We found many SNPs (n = 153) with high iHS scores (|iHS| > 2) in different ethnic groups in a 78 kb region encompassing the *TMPRSS2* gene which show high levels of LD (ChrX:41454000-41541000; Figure S16, S17). We identified a non-synonymous variant (rs150969307) that shows a signature of positive selection (iHS = 2.01) and is common only in the Chabu hunter gatherer population from Ethiopia (MAF = 0.079) (Table S11). We found that more than one third of SNPs with |iHS| scores >2.0 (62 of 153) are located in putative regulatory regions (Figure S18 and Table S8).

### Associations between genetic variations in *TMPRSS2* and clinical disease phenotypes

In the PMBB, gene-based analysis with 12 severe disease classes identified nominal associations with respiratory failure, respiratory syncytial virus infectious disease, lower respiratory tract infection and pneumonia in the AA population but no statistically significant association in the EA population (Table 1, Figure 4E). As with *ACE2*, the clinical phenotype associations with *TMPRSS2* in AA may be driven by an excess of rare variants in that population and, hence, more carriers in comparison to EA. Among the diseases in the respiratory disorder category, we identified a nominal association (p<0.01) with allergic rhinitis in AA and with obstructive bronchitis in EA populations (Table S9). Previously, a gene-based PheWAS with EHR-derived disease codes in the UK biobank population, which consists of mostly individuals of European descent, showed no statistically significant associations with *TMPRSS2*^55^. Among clinical lab measures, we identified nominal association with urine bilirubin levels (p = 0.001). The PheWAS of the two regulatory eQTLs (rs76833541 and rs4283504) of *TMPRSS2* described above identified association of rs76833541 with abnormal glucose (p=8.9 x 10^-4^, OR=1.5) in EA and rs4283504 with glucocorticoid deficiency (p= 0.001, OR=2.7) in AA (Figure 4F). We did not identify any association between these two eQTLs and respiratory conditions (Figure 4F, and Table S10).

### Patterns of variation at *DPP4* and *LY6E*

#### DPP4

*DPP4* is a receptor for the Middle East Respiratory Coronavirus (MERS-Cov) and was reported to interact with SARS-CoV-2^56^. At this gene, we identified 47 non-synonymous variants and one loss-of-function variant (Table S12). Among them, no variant was common in the pooled global dataset (Figure 6A), suggesting this gene is extremely conserved during human evolutionary history. Only one non-synonymous variant (rs1129599, Ser437Thr) was common in the Fulani pastoralists from Cameroon (MAF = 0.081), was present at low frequency in other African populations, and was absent in non-African populations (Figure 6B and 6C). In addition to the nonsynonymous variants, one loss-of-function variant was identified at *DPP4*. The variant rs149291595 (Q170*) has low MAF in some African populations (MAF < 0.05) but is absent in non-African populations.

**Figure 6.**
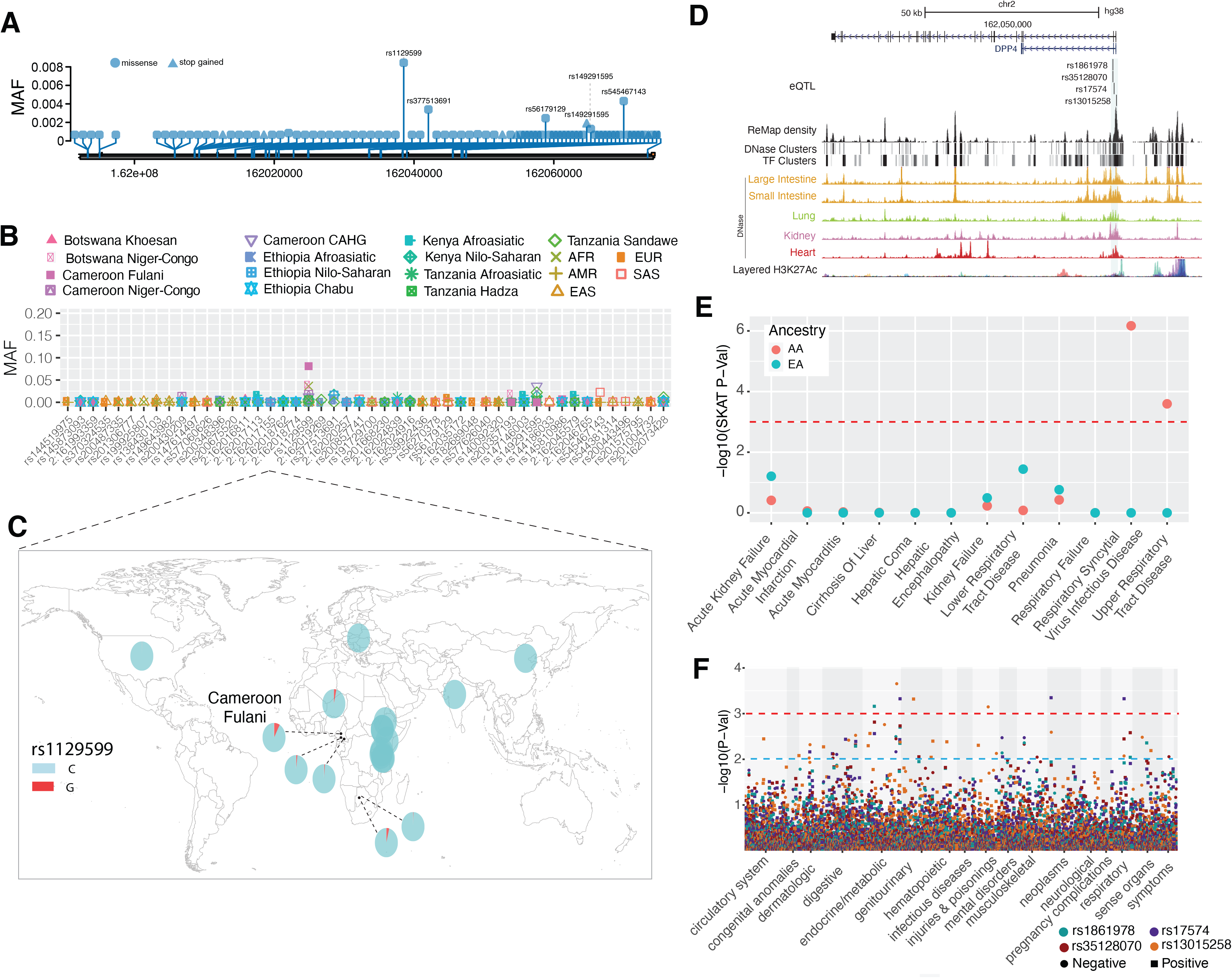
Genetic variation at *DPP4* and its disease association. (A) Location of coding variants and their minor allele frequency (MAF) at *DPP4* identified from the pooled dataset. (B) MAF of coding variants in diverse global ethnic groups. (C) The MAF of variant within rs129559 at *DPP4* in diverse global ethnic groups. (D) Regulatory eQTLs located in *DPP4*. RNA Pol2 ChIA-PET data and DNase-seq data of large intestine, small intestine, lung, kidney and heart are from ENCODE ^81^. (E) Gene-based association result between coding variants at *DPP4* and 12 disease classes. The disease classes are shown on the x-axis and the y-axis represents the p-values. EA, European Ancestry; AA, African American ancestry. (F) PheWAS plot of the four eQTLs associated with *DPP4* and ∼1800 disease codes across 17 disease categories. The disease categories are shown on the x-axis and the y-axis represents the -log10 of the p-values. The colored dot represents an eQTL and the direction of effect of the association. The red dashed line denotes the 0.0001 cutoff, and the blue dashed line represent the 0.001 cutoff.

We identified four eQTLs (rs1861978, rs35128070, rs17574 and rs13015258) in the promoter region of the *DPP4* gene (Figure 6D). Three of the variants (rs1861978, rs35128070 and rs17574) are significant eQTLs in the transverse colon and rs13015258 is an eQTL in the lung (P < 5.9e-6, Figure S20 and Table S4). The minor alleles of these three variants are rare in EAS (MAF < 0.05) but common in all other populations (MAF > 0.15, Figure S21, and Table S2). The fourth SNp, rs13015258, resides in the center of a cluster of DNase peaks identified in ENCODE (Figure 6D) with MAF ranging from 0.38 in the AMR population to 0.6 in other populations (Figure S21 and Table S2).

### Signatures of natural selection at *DPP4*

The MK-test result was not significant in either the pooled samples (Dn = 3, Ds = 5, Pn = 45, Ps = 33 OR = 0.44, P = 0.9, two-sided Fisher’s exact test) nor in each population separately (Table S6 and Figure S8). For the the dN/dS test, we observed ratios ranging from 0 to 0.52 in individual populations, indicating that *DPP4* is highly conserved (Table S5 and Figure S6) within human populations. Using the iHS test, we identified 8 SNPs that had extreme high iHS scores (|iHS| > 2) in the Khoesan populations from Botswana (Figure S22 and Table S7). Five of these SNPs (rs10166124, rs2284872, rs2284870, rs7608798 and rs2160927) are in LD (D’ > 0.95) with each other (Figure S23). The SNP rs2284870 is located in a strong DNase peak in heart tissue (Figure S24 and Table S8).

### Associations between genetic variations in *DPP4* and clinical disease phenotypes

In the gene-based analysis among AA PMBB participants, we identified significant associations (only in the SKAT model) with respiratory syncytial virus infectious disease and upper respiratory tract disease (Figure 6E, Table 1 and S9). None of the gene-based models were significant in the EA population. The PheWAS of four regulatory eQTLs identified the most significant association with malignant neoplasm of the rectum (commonly referred as colon cancer) for rs17574 (p = 4.49 x 10^-04^, OR = 1.8) and rs13015258 (p = 0.002, OR = 0.54) in AFR only. Among respiratory disorders, rs35128070 had the most significant association with “abnormal results of function study of pulmonary system” (p=0.002, OR=1.6) in the AFR population and we observed a nominal association between rs17574 and “acute respiratory infections” (p<0.01, OR=1.22) in the EA population (Figure 6F, and Table S10).

#### LY6E

Studies show that mice lacking *LY6E* were highly susceptible to a usually nonlethal mouse coronavirus^24^. At *LY6E* we observed twenty-eight non-synonymous variants and all of them, except rs11547127 (MAF = 0.057), have MAF that are rare in the pooled global dataset (Figure 7A, and Table S13). However, some of the non-synonymous variants are common in specific populations (Figure 7B). For instance, the non-synonymous variant rs111560737 (Asp104Asn) was common in the southern African Khoesan population from Botswana (MAF = 0.36) and the Chabu population from Ethiopia (MAF = 0.17) (Figure 7C). Three loss-of-function variants (rs200177123 [stop gained, Ser59*], chr8:143020941, and chr8:143020946) were also identified at *LY6E*, and all of them are rare. In the PMBB, only four pathogenic and likely pathogenic variants were identified, and all were rare in both AA and European EA populations.

**Figure 7.**
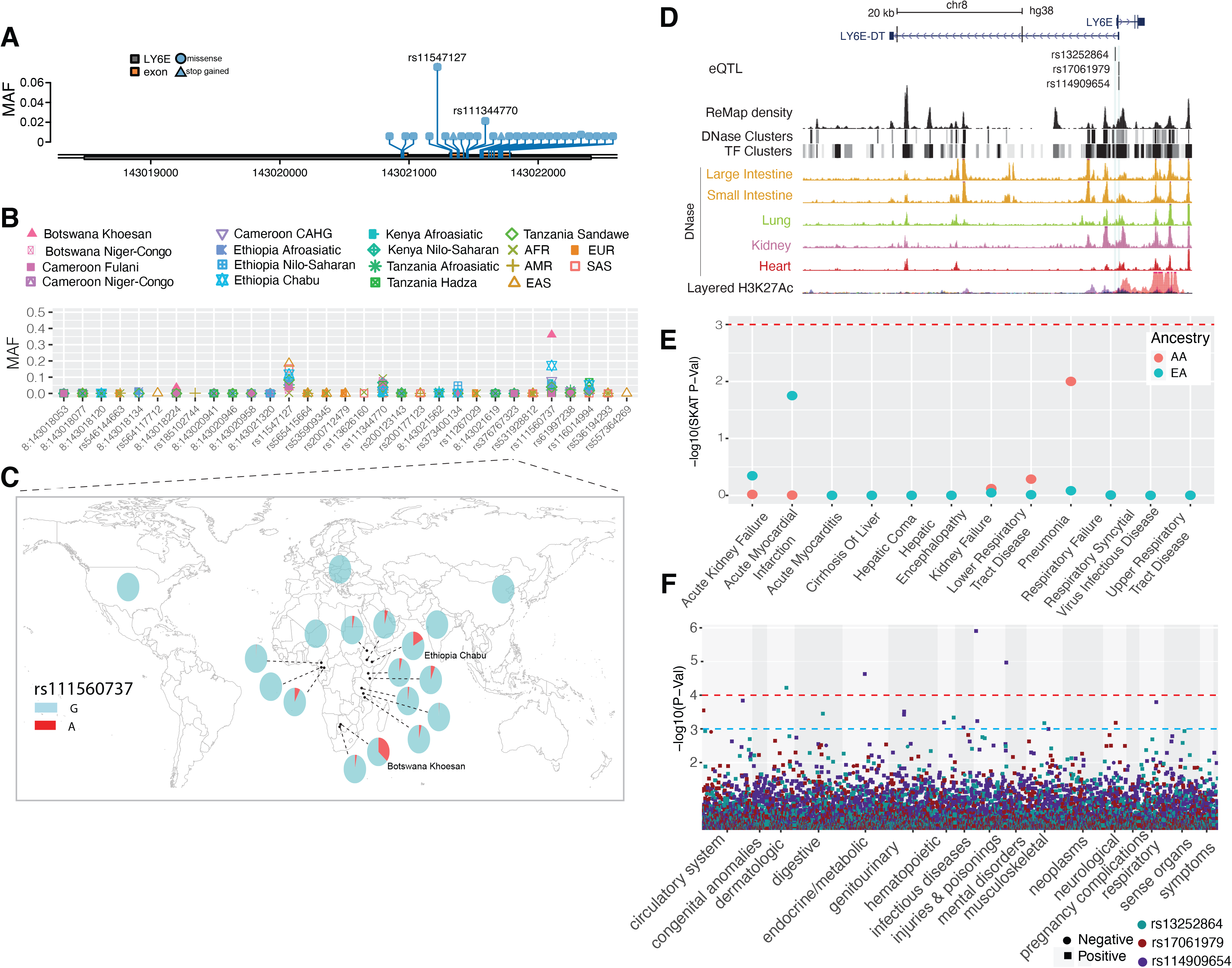
Genetic variation at *LY6E* and its disease association. (A) Location of coding variants and their minor allele frequency (MAF) at *LY6E* identified from the pooled dataset. (B) MAF of coding variants in diverse global ethnic groups. (C) The MAF of variant rs111560737 at *LY6E* in diverse global ethnic groups. Each pie denotes frequencies of alleles in the corresponding population. (D) Three regulatory eQTLs identified at *LY6E*. RNA Pol2 ChIA-PET data and DNase-seq data of large intestine, small intestine, lung, kidney and heart are from ENCODE ^81^. (E) Gene-based association result between coding variants at *LY6E* and 12 disease classes. The disease classes are shown on the x-axis and the y-axis represents the p-values. EA, European Ancestry; AA, African American ancestry. (F) PheWAS plot of the three eQTL associated with *LY6E* and ∼1800 disease codes across 17 disease categories. The disease categories are shown on the x-axis and the y-axis represents the -log10 of the p-values. The colored dot represents an eQTL and the direction of effect of the association. The red dashed line denotes the 0.0001 cutoff, and the blue dashed line represents the 0.001 cutoff.

We identified three regulatory eQTLs (rs13252864, rs17061979 and rs114909654) located within 2 kb of the transcription start site of *LY6E* (Figure 7D), all of which are significant in esophageal mucosa (P < 1e-5, Figure S25 and Table S4), which has a high expression level of *LY6E* (TPM=108, GTEx). The minor alleles of rs13252864 and rs114909654 are common in African populations (MAF > 0.15) while very rare in other populations (MAF < 0.02, Figure S26), whereas the MAF of rs17061979 is relatively high in EAS (0.18) and SAS (0.13) and rare in other populations (MAF < 0.05, Figure S26).

### Signatures of natural selection at *LY6E*

The MK-test result was not significant in either the pooled samples (Dn = 0, Ds = 4, Pn = 9, Ps =9, OR = 0, P = 0.9, two-sided Fisher’s exact test) nor in each population separately (OR ranging from 0 to 0.52; Table S6 and Figure S8), indicating that *LY6E* is highly conserved. We identified 19 variants that that had extreme high iHS scores (|iHS| > 2) (Table S7, Figure S27), some of which are in LD in specific populations (Figure S28). One variant (rs867069115) shows an extreme iHS score in the Hadza hunter-gatherer population from Tanzania (iHS = -2.94). This variant is located in a regulatory region ∼1.9kb downstream of *LY6E*, within DNase and TF peaks in the lung, intestine, kidney, heart, stomach, pancreas and skeletal muscle from ENCODE (Figure S29) and is common only in the Hadza population (MAF = 0.14), is rare in other African population (MAF < 0.05) and is absent in all non-African populations (Table S2). SNP rs10283236, which shows an extreme iHS value in the CEU population, is an eQTL of *LY6E* located within DNase and TF clusters identified in ENCODE (∼4.14kb downstream of *LY6E*) active in many tissues including lung, kidney and small intestine.

### Associations between genetic variations in *LY6E* and clinical disease phenotypes

We identified a nominal association between *LY6E* with pneumonia in the AA population only (p= 0.01, Figure 7E, Table 2 and Table S9). *LY6E* also has nominal association (p<0.01, Table 2) with total cholesterol, prothrombin, and eosinophil levels among the AA population. Association with prothrombin is not statistically significant in the EA population. The association analysis of regulatory variants identified most significant association with “severe protein-calorie malnutrition” (p = 2.35 x 10^-05^, OR = 1.9) and “acute post hemorrhagic anemia” (p = 6.4 x 10^-04^, OR = 1.6) in the AA population. In the EA population, “chronic ulcer of skin” with rs13252864 (p=0.001, OR=2.2) was the most significant association (Figure 7F, and Table S10).

## Discussion

Investigating global patterns of genetic variation at genes that play a role in SARS-CoV-2 infection could provide insights into potential differences in susceptibility to COVID-19 among diverse human populations. However, African populations are under-represented in the majority of current genetic studies of COVID-19 susceptibility and severity, despite the fact that they have the highest genetic diversity among human populations^57; 58^. In this study, we present a comprehensive analysis of human genes which play a key role in SARS-CoV-2 host receptor binding and cellular invasion, i.e., *ACE2, TMPRSS2, DPP4,* and *LY6E*. We characterized the coding and non-coding variants in these candidate genes to examine population differences in allele frequencies and signatures of natural selection in diverse ethnic populations. This included novel sequence data from 2012 ethnically diverse African populations from five countries (Cameroon, Ethiopia, Kenya, Botswana and Tanzania) in Africa practicing different lifestyles (e.g. hunter-gatherers, agriculturalists, and pastoralists). Additionally, we analyzed the correlation of common and rare genetic variants in these four genes with clinical traits derived from the dataset of 15,997 individuals from the Penn Medicine BioBank (PMBB) with African and European ancestry. We included 12 “organ dysfunction” categories defined by phenotype algorithms (see Methods), ∼1800 ICD diagnosis codes, and 33 laboratory test measures from the EHR. Our results highlight the importance of including genomes from diverse ethnic groups in human genetic studies.

At *ACE2* we identified 41 non-synonymous variants, most of which are rare, suggesting that they are under purifying selection. Tests based on dN/dS indicate that East Asians have an excess of non-synonymous variation at *ACE2*, indicating weak purifying selection has influenced patterns of variation in that population. However, there are some variants that are common in specific ancestry groups. Notably, we identified three common non-synonymous variants (rs138390800, rs147311723, and rs145437639) at *ACE2* with MAF ranging from 0.083 to 0.164 in Central African hunter-gatherers (CAHG), which were the only common coding variants (defined here as MAF > 0.05) found in global populations studied here and by others^20; 53; 59; 60^. We observed that the derived alleles of the common non-synonymous SNPs (rs138390800, rs147311723, rs145437639) and one putative regulatory variant (rs186029035) at *ACE2* in *CAHG* show evidence of EHH, with the extended haplotypes extending longer than 2 Mb, though they did not show deviation from neutrality based on the iHS test. However, we do not have much power to detect a selection signal using this test because the SNPs are on three different haplotype backgrounds in CAHG, possibly due to selection on existing variation (e.g.“soft selection”) which decreases the power to detect significant iHS scores. Moreover, each haplotype is at a relatively low frequency (0.083 to 0.164), which further reduces the power of the iHS test. The CAHG are traditionally hunter-gatherers living in a rainforest ecosystem who consume wild animals. They have high exposure to animal viruses and were reported to have relative resistance to viral infection^61^. Thus, it is possible that this locus is adaptive for protection from infectious diseases in this population. Future *in vitro* or *in vivo* studies will be needed to determine the functional significance of these variants.

At *TMPRSS2*, we identified forty-eight nonsynonymous variants, only two of which had a high MAF (>.05) in the pooled global dataset (rs12329760 and rs75603675). However, some variants have high MAF in two African hunter-gatherer populations. Notably, the non- synonymous variant rs61735795 (Pro375Se) is only common in the Khoesan-speaking San population from Botswana (MAF = 0.18) and the non-synonymous variant rs367866934 (Leu403Phe) is only common in the Cameroonian CAHG populations (MAF = 0.15). At *TMPRSS2* we observed a strong signature of adaptive evolution in the human lineage after divergence from Chimpanzee ∼ 6 MYA^62^. In total, 13 non-synonymous variants located on different structural domains of TMPRSS2 were fixed in human populations. Among them, E441Q and T515M are located in the Peptidase S1 domain that plays an important role in acute respiratory syndrome (SARS)-like coronavirus (SARS-CoV-2) infection^63^ and six (A3P, N10S, T46P, A70V, R103C, and M104T) are at the cytoplasmic amino terminal domains of *TMPRSS2* which plays an important role in signal transduction. These variants at *TMPRSS2* could be potential candidates for future studies to investigate their functional impact on susceptibility to pathogens in humans compared to non-human primates.

SARS-CoV replication is significantly reduced in *ACE2* knockout mice^64^ and cells with low expression of *ACE2* were resistant to SARS-CoV2 infection^65^. It has also been shown that both SARS-CoV and SARS-CoV2 infection could down regulate *ACE2* expression^22; 64; 66^. The expression of *ACE2* and *TMPRSS2* in nasal and bronchial epithelial cells is higher in adults than children, and in healthy individuals compared with smokers or patients with chronic obstructive pulmonary disease^51^. Therefore, differences in expression levels of *ACE2* and *TMPRSS2* could influence the susceptibility and host reactions to SARS-CoV-2. Regulatory eQTLs that differ in frequency across ethnically diverse populations may play a role in local adaptation and disease susceptibility^67^. eQTL mapping has been used to identify population-specific regulatory variation and revealed the association of regulatory alleles with complex traits such as multiple sclerosis^68^, malaria^54^ and immune response to infection^69^. We identified regulatory eQTLs associated with *ACE2*, *TMPRSS2*, *DPP4*, and *LY6E* gene expression and highlighted the eQTLs showing highly differentiated MAF among populations and/or signatures of natural selection. These eQTLs are located in ChIP-seq and DNase peaks and have the potential to influence transcription factor binding and, thus, change the promoter or enhancer activities in specific tissues^70; 71^. Interestingly, some of the eQTLs in the upstream regions of *ACE2* were under selection in African populations. For example, rs5936010 and rs5934263, which are located within a strong enhancer interacting with the promoter of *ACE2* as suggested by ChIA-PET, harbored significant iHS scores (|iHS| > 2) in both Afroasiatic populations from Kenya and the San population from Botswana. Further, PheWAS of these eQTLs in the PMBB populations identified association of eQTLs at *ACE2* with type 2 diabetes (rs5936010) and hypertension (rs5934263). These are known pre-existing conditions that increases risk of severe illness due to COVID-19^11; 72; 73^. Among respiratory diseases, only one eQTL at *ACE2* had nominal association (rs4830977) with acute sinusitis. The association was only identified in the AA population and had a protective effect (OR = 0.78 [0.66-0.95]). The eQTLs we analyzed are from GTEx V8 database^74^, and 84.6% of the donors are people of European and Western Eurasian descent. Therefore, it is possible that we are missing some regulatory variants that are only present in specific ancestry groups due to the lack of sample diversity. Further experimental testing of predicted regulatory variants will provide insights into differences in gene expression regulation at *ACE2*, *TMPRSS2*, *DPP4*, and *LY6E* among different populations. In the future, eQTL mapping in diverse populations will be informative for identifying novel trait associations that may differ in prevalence across ethnic groups^75^.

The gene-based genetic association analyses of non-synonymous variants at *ACE2*, *TMPRSS2*, *DPP4* and *LY6E* identified several associations with clinical phenotypes. We observed that respiratory failure has significant association with *ACE2* and *TMPRSS2* among the PMBB AA population. That is a particularly interesting finding as respiratory failure is one of the clinical outcomes observed in some patients with COVID-19^10; 45–48^. However, this association was not significant in the EA population. This observation could be explained by the low number of coding variants and carriers at *ACE2* and *TMPRSS2* among EA and, hence, low power to detect an association. An association with myocarditis, a rare cardiovascular disease caused by viral infection, was also observed in the AA population. Recent studies have reported a link between SARS-CoV-2 induced cardiac injury such as myocarditis among COVID-19 patients^76^. Further, *ACE2* has known expression in heart tissue, and it plays an important role in transcriptional dysregulation in cardiomyocytes – cells that make up cardiac muscles^50–52^. We observed association between ACE2 and myocarditis only in the AA population but as noted above, we may not have as much power to detect and association in EA. Blood clotting abnormalities in lungs and other organs in COVID-19 patients have been reported by several studies^77^. In autopsies of COVID-19 patients, thrombosis was found to be a prominent finding across multiple organs, even in spite of extensive anticoagulation treatment and regardless of timing of clinical progression, indicating that thrombosis might be at play in the early stages of disease^77^. One hypothesis to explain this observation is that the dysfunction of endothelial cells may play an important role in increased risk of thrombosis^78^. We observed associations between the internationalized normalized ratio (INR) derived from the prothrombin time test (PT) with *ACE2* and *LY6E* in a gene-based association test. The INR test measures the time it takes blood to clot and is an important measure for individuals with blood clotting disorders or on blood thinners.

Characterizing the genetic variation and clinical phenotype associations at these four genes that play a key role in SARS-CoV-2 infection could be relevant for understanding individual and population differences in infection susceptibility. We performed evolutionary analyses to dissect the forces underlying global patterns of genetic variation and identified variants that may be targets of selection. It will be important to determine the functional effects of these candidate adaptive variants using *in vitro* and in *vivo* approaches in future studies. Additional studies will be needed to investigate the impact of genetic variation in modulating susceptibility/resistance to SARS-CoV-2 infection and other coronaviruses across ethnically diverse populations.

## Material and Methods

### Genomic data

The genomic data used in this study were from three sources: the Africa 6K project (referred to as the the “African Diversity” dataset) which is part of the TopMed consortium^79^, the 1000 Genomes project (1KG)^25^, and the Penn Medicine BioBank (PMBB). From the Africa 6K project, a subset of 2012 high coverage (>30X) whole genome sequences of ethnically diverse African populations (Figure S1) were included. The African samples were collected from individuals from five countries (Cameroon, Ethiopia, Kenya, Botswana and Tanzania), speak languages belonging to four different language families spoken in Africa (Afroasiatic, Nilo- Saharan, Niger-Congo, and Khoesan) and have diverse subsistence practices (*e.g.,* hunter- gatherers, agriculturalists, and pastoralists). IRB approval was obtained from the University of Maryland and the University of Pennsylvania. Written informed consent was obtained from all participants and research/ethics approval and permits were obtained from the following institutions prior to sample collection: COSTECH, NIMR and Muhimbili University of Health and Allied Sciences in Dar es Salaam, Tanzania; the University of Botswana and the Ministry of Health in Gaborone, Botswana; the University of Addis Ababa and the Federal Democratic Republic of Ethiopia Ministry of Science and Technology National Health Research Ethics Review Committee; and the Cameroonian National Ethics Committee and the Cameroonian Ministry of Public Health. Whole genome sequencing (WGS) was performed to a median depth of 30X using DNA isolated from blood, PCR-free library construction and Illumina HiSeq X technology, as described elsewhere^79^. In the 1KG data set, 2504 genome sequences from phase 3^25^ were included in our analysis.

The PMBB participants were recruited through the University of Pennsylvania Health System by enrolling at the time of clinic visit. Patients participate by donating either blood or a tissue sample and allowing researchers access to their EHR information. This academic biobank has DNA extracted from blood that has been genotyped using an Illumina Infinium Global Screening Array-24 Kit *version 2* and whole exome sequencing (WES) using the IDT xgen exome research panel v1.0. The study cohort consisted of 15,977 individuals total, with 7,061 of European ancestry (EA) and 8,916 of African ancestry (AA) (Table S1). Genetic ancestry of these samples was determined by performing quantitative discriminant analyses (QDA) on eigenvectors. The 1000 Genomes datasets with super population ancestry labels (EUR, AFR, EAS, SAS, Other) were used as QDA training datasets to determine the genetic ancestry labels for the PMBB population. We identified and removed 117 related individuals using a kinship coefficient of 0.25.

### Variant annotations

We used Ensembl Variant Effect Predictor (VEP) for variant annotations^80^. VEP classifies variants into 36 types including non-synonymous, synonymous, and stop loss variants. For pathogenicity predictions, we used CADD^26^, SIFT^27^, PolyPhen^28^ , Condel^29^, and REVEL scores in Ensembl. For whole-genome sequencing datasets (African Diversity and 1KG), we annotated genetic variants at *ACE2* (chrX:15,561,033-15,602,158), *TMPRSS2* (chr21:41,464,305-41,531,116), *DPP4* (chr2:161,992,245-162,074,215) and *LY6E* (chr8:143,017,982-143,023,832), and 10 Mb flanking these genes (Table S2). For whole-exome genomes from the PMBB dataset, annotations were restricted to coding regions only. For gene-based association analysis using the PMBB dataset, we collapsed all the predicted non-synonymous variants with REVEL score > 0.5 and putative loss of function variants (pLOFs) with MAF < 0.01. We assigned variants as pLoFs if the variant was annotated as stop_lost, missense_variant, start_lost, splice_donor_variant, inframe_deletion, frameshift_variant, splice_acceptor_variant, stop_gained, or inframe_insertion. All genome coordinates followed the GRCh38 assembly.

### Characterization of putative regulatory variation

We identified regulatory variants likely to impact the target genes. For all four genes (*ACE2*, *TMPRSS2*, *DPP4* or *LY6E*), we extracted the variants located within ±10kb distance to their TSS as well as enhancers supported by RNA Pol2 ChIA-PET data from ENCODE^81^. These variants were further filtered by overlapping with DNase-seq and ChIP-seq peaks from Roadmap^82^, ENCODE^81^, Remap2 ^83^; or overlapping with significant single-tissue expression quantitative trait locus (eQTLs) (P-value<0.001) from the GTEx V8 database ^32^. We visualized the location of these regulatory and eQTL variants using the UCSC genome browser and highlighted the variants using Adobe Illustrator.

### Electronic Health Record Phenotypes

In this analysis, we focused on the phenotypes characterized as primary organ dysfunctions in the early studies on COVID-19. Broadly, we centered our analyses on these four broad clinical conditions/phenotypes: respiratory injury/failure, acute liver injury/failure, acute cardiac injury/failure, and acute kidney injury/failure. These disease classes are well characterized in human disease ontologies such as Monarch Disease Ontology (MONDO). MONDO merges multiple disease resources such as SNOMED, ICD-9, and ICD-10. We leveraged the existing mappings between ICD-9/10 codes (which are how the data are coded in the EHR) and the MONDO disease classes for the conditions described above. We identified 12 MONDO classes that are closely related to four conditions of interest (Table S1). By using ICD- 9 and ICD-10 data from the EHR of the PMBB participants, we mapped the ICD codes to 12 MONDO disease classes. Details on the ICD code mapping to MONDO disease classes are provided in Table S14. Individuals were defined as cases if they had at least one instance of any ICD code mapped to a MONDO disease class or as controls if they had no instance of the code in that disease class. A clinical expert on our team manually reviewed the MONDO and ICD- 9/10 mappings.

We also used EHR phenotypes defined by groupings of ICD-9 and ICD-10 codes into clinically relevant groups, called phecodes, used in prior PheWAS studies^84^. Individuals with two or more instances of a phecode were defined as cases, whereas those with no instance of a phecode were defined as controls. Individuals with only one instance were excluded for that phecode. A total of 1860 phecodes were included in the study.

Additionally, we extracted data on 34 clinical laboratory measures for PMBB participants from the EHRs. We derived a median value for each laboratory measure based on all clinical tests ever done within the Penn Medicine health system. Any measurement value that falls more than three standard deviations from the normal were labeled as outliers and removed.

### Association Testing

We used the R SKAT package for conducting a gene-based dispersion test and Biobin^42; 85^ for gene burden analysis. Here, multiple genetic variations in a gene region were collapsed to generate a gene burden/dispersion score and regression methods were used to test for association between the genetic score and a phenotype or trait. We performed three separate burden analysis for 12 MONDO disease classes (Table S14), 1860 phecode, and 34 clinical lab measures. Briefly, the variants annotated as non-synonymous (REVEL score >= 0.5) and pLoFs within each of the four candidate genes were collapsed into their respective gene regions *(ACE2, TMPRSS2, DPP4 and LY6E).* For both statistical dispersion and burden tests, models were adjusted by the first four principal components of ancestry, sex, and decade of birth. For multiple hypothesis correction, a conservative Bonferroni adjustment was used to derive a significant p-value threshold (p-value < 0.0001). We also performed a univariate statistical test for each of the rare variants from these four candidate gene regions to study the effects of each single nucleotide variant (SNV) on the disease phenotype.

### Structural analysis of nonsynonymous variations on ACE2-S protein binding interface

The fast response from the structural biology community to the COVID-19 pandemic led to the exceptionally fast determination and publication of over 900 as of Jan. 2021 (https://www.rcsb.org/news?year=2020&article=5e74d55d2d410731e9944f52&feature=true) protein structures related to SARS-Cov-2. Using experimentally determined structures of the ACE2 protein complexed with the receptor binding domain (RBD) of SARS-CoV-2 spike glycoprotein, we assessed possible impacts of nonsynonymous coding variants on the ACE2- binding interface with SARS-CoV-2-RBD. Among the multiple entries available in the Protein Data Bank (PDB), we chose to focus on the structure of the full-length human ACE2 bound to RBD (PDB ID 6M17 ^86^) determined with Cryo-Electron Microscopy (cryo-EM), as it presented multiple advantages to our study. Unlike other PDB entries that only feature sections of ACE2, usually focusing on the part of the enzymatic domain responsible for RBD binding, 6M17 presents the full length ACE2 in its dimeric form. This allowed us to identify the 3D protein location of all nonsynonymous coding variants identified in this study. Moreover, ACE2 was expressed in a human cell line, maintaining important glycosylation sites and allowing the cryo- EM structure to be used to identify their positions and compositions ^86^. All structural analysis and figures were prepared using VMD ^87^.

### Detecting signatures of natural selection

We used two methods (the McDonald–Kreitman test ^35^ and the Dn/Ds test ^34^) to test for signals of selection acting on the four candidate genes over long time scales, and two methods (EHH and iHS) to detect recent (e.g. last ∼10,000 years before present) signatures of positive selection .

For the McDonald–Kreitman test (MK-test) ^35^, we set up a two-way contingency table to statistically compare the number of nonsynonymous (Dn) and synonymous (Ds) fixed differences between humans and chimpanzees with the number of nonsynonymous (Pn) and synonymous (Ps) polymorphisms among individuals within a population. Based on neutral theory, the ratio of nonsynonymous to synonymous changes should be constant throughout evolutionary time, i.e. the ratio observed among individuals within species (Pn/Ps) should be equal to the ratio observed between species (Dn/Ds). Under a hypothesis of positive selection in the hominin lineage after divergence from our closest ancestor, the chimpanzee, the ratio of nonsynonymous to synonymous variation within species is expected be larger than the ratio of nonsynonymous to synonymous variation between species (i.e. Dn/Ds > Pn/Ps). If there is positive diversifying selection among human populations but conservation of fixed differences between species, the ratio of nonsynonymous to synonymous variation between species should be lower than the ratio of nonsynonymous to synonymous variation within species (i.e. Dn/Ds < Pn/Ps). The chimpanzee sequence (Clint_PTRv2/panTro6) used in the analysis was obtained from the UCSC genome browser. We used Fisher’s exact test to detect significance of the MK- test. We used transcripts ENST00000252519.8, ENST00000398585.7, ENST00000360534.8, ENST00000521003.5 to calculate Dn, Ds, Pn and Ps for *ACE2*, *TMPRSS2*, *DPP4* and *LY6E*, respectively.

We also used the ratio of substitution rates at non-synonymous and synonymous sites (dN/dS) to infer selection pressures on the four candidate genes, as the dN/dS ratio has more power to detect recurrent positive selection^88^. This measure quantifies selection pressures by comparing the rate of substitutions at synonymous sites (dS), which are neutral or close to neutral, to the rate of substitutions at non-synonymous sites (dN), which are more likely to experience selection. The dN/dS estimation used here follows Nei et al^34^. The number of synonymous sites, s, for codon i in one protein is given by

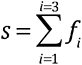

where *f*_i_ is defined as the proportion of synonymous changes at the *i*th position of a codon. For a sequence of *r* codons, the total number of synonymous sites, *S* is given by

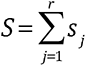

where s_j_ is the value of *s* at the *j*th codon, and the total number of non-synonymous sites, *N = 3r - S*. The total number of synonymous and non-synonymous differences between two sequences, *S_d_* and *N_d_* respectively, are given by

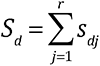

and

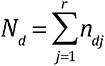

where *s_dj_* and *n_dj_* are the numbers of synonymous and non-synonymous differences between two sequences for the *j*th codon, and *r* is the number of codons compared. The proportions of synonymous (*pS*) and non-synonymous (*pN*) differences are estimated by the equations *pS = Sd / S* and *pN = Nd / N*. The numbers of synonymous (*dS*) and non- synonymous (*dN*) substitutions per site are estimated using the Jukes-Cantor formula as below:

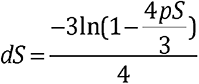

and

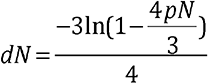

In our analysis, for each population, we estimated the total number of synonymous (*S_d_*) and non- synonymous (*N_d_*) differences, and then calculated *dN*/*dS*. If *dN*/*dS* is larger than one, it suggests positive diversifying selection influencing variation at the gene. If *dN*/*dS* is less than one it suggests the gene is evolutionary conserved.

Genomic regions that have undergone recent positive selection are characterized by extensive linkage disequilibrium (LD) on haplotypes containing the mutation under selection. We used the extended haplotype homozygosity (EHH)^39^ and the integrated Haplotype Score (iHS) methods^38^ to identify regions with extended haplotype homozygosity greater than expected under a neutral model. iHS is based on the differential levels of LD surrounding a positively selected allele compared to the ancestral allele at the same position. For the iHS analyses, we normalized scores with respect to all values observed at sites with a similar derived allele frequency within 40Mb regions flanking the four target genes. SNPs with absolute values larger than 2 are within the top 1% of observed values and are marked as extreme SNPs or candidate SNPs under positive selection. An extreme positive iHS score (iHS > 2) means that haplotypes on the ancestral allele background are longer compared to the derived allele background. An extreme negative iHS score (iHS < -2) means that the haplotypes on the derived allele background are longer compared to the haplotypes associated with the ancestral allele. All of the above processes were performed with selscan^89^. SNPs with predicted functional effects on protein structure that are identified as potential targets of selection (stop_lost, missense_variant, start_lost, splice_donor_variant, inframe_deletion, frameshift_variant, splice_acceptor_variant, stop_gained, or inframe_insertion) are highlighted. Haplotypes were phased by Eagle V2.4.1^90^. The ancestral state of alleles was obtained from Ensembl.

To identify potential regulatory variants under selection, we overlapped SNPs showing signatures of selection using iHS with DNase I hypersensitivity peak clusters from ENCODE ^81^ and eQTLs from GTEx v8. ^32^. The overlapped SNPs were uploaded to the UCSC browser for visualization. The ChIP-seq density dataset was obtained from http://remap.univ-amu.fr/ ^82^. DNase-seq and ChIP-seq clusters, layered H3K4Me3 (often found near Promoters), H3K4Me1 and H3K27Ac (often found near Regulatory Elements) data are from ENCODE^81^. The DNase-seq tracks of large intestine, small intestine, lung, kidney, heart, stomach, pancreas and skeletal muscle were from ENCODE ^91^.

We used *d_i_* statistics to identify SNPs that are highly differentiated in allele frequency between populations based on unbiased estimates of pairwise F*_ST_* ^41^. The *d_i_* statistics were performed cross the 40Mb regions. If the candidate SNP was within the top 5% of the 40Mb regions in a specific population, the SNP was considered as a variant showing significant differentiation between the target population and other populations. These variants are candidate SNPs that show signals of local adaptation.

Haplotype networks were constructed by PopART^92^ using the built-in minimum spanning algorithm.

## Description of Supplemental Data

Supplemental file 1: Supplemental figures S1-S29.

Table S1. Penn Medicine Biobank (PMBB) participant characteristics

Table S2. Genetic variants identified around the four genes. “N” denotes variants were not identified or called in the corresponding dataset. “0” denotes variants were identified in the corresponding dataset, but the minor allele frequency is 0.

Table S3. Coding variants identified at *ACE2*. “N” denotes variants were not identified or called in the corresponding dataset. “0” denotes variants were identified in the corresponding dataset, but the minor allele frequency is 0.

Table S4. Regulatory variants identified at the four candidate genes. eQTLs are extracted from GTEx V8.

Table S5. Result of the dN/dS for four genes in both the pooled dataset and specific ethnic groups.

Table S6. Results of the MK-test for four genes in both the pooled dataset and specific ethnic groups.

Table S7. SNPs with significant selection signals in each ethnic group based on each method.

Table S8. Regulatory SNPs that overlap with significant selection signals at the four genes.

Table S9. Summary statistics from gene-based association results

Table S10. Summary statistics from PheWAS of eQTL variants

Table S11. Coding variants identified at *TMPRSS2*. “N” denotes variants were not identified or called in the corresponding dataset. “0” denotes variants were identified in the corresponding dataset, but the minor allele frequency is 0.

Table S12. Coding variants identified at *DPP4*. “N” denotes variants were not identified or called in the corresponding dataset. “0” denotes variants were identified in the corresponding dataset, but the minor allele frequency is 0.

Table S13. Coding variants identified at *LY6E*. “N” denotes variants were not identified or called in the corresponding dataset. “0” denotes variants were identified in the corresponding dataset, but the minor allele frequency is 0.

Table S14. ICD code mapping to MONDO disease classes.

## Data Availability

Additional information for reproducing the results described in the article is available upon reasonable request and subject to a data use agreement.

## Acknowledgements

Molecular data for the Trans-Omics in Precision Medicine (TOPMed) program was supported by the National Heart, Lung and Blood Institute (NHLBI). Genome Sequencing for “NHLBI TOPMed: Integrative Genomic Studies of Heart and Blood Related Traits in Africans (Africa6K)” was performed at Broad Genomics (hhsn268201600034i). Core support including centralized genomic read mapping and genotype calling, along with variant quality metrics and filtering were provided by the TOPMed Informatics Research Center (3R01HL-117626-02S1; contract HHSN268201800002I). Core support including phenotype harmonization, data management, sample-identity QC, and general program coordination were provided by the TOPMed Data Coordinating Center (R01HL-120393; U01HL-120393; contract HHSN268201800001I). We gratefully acknowledge the studies and participants who provided biological samples and data for TOPMed.

Funding for this study was provided by grant numbers: X01HL139409, 1R35GM134957, R01GM113657, R01DK104339, ADA 1-19-VSN-02, R01AR076241 to SAT, and R01LM010098 to SW. Cesar de la Fuente-Nunez holds a Presidential Professorship at the University of Pennsylvania, is a recipient of the Langer Prize by the AIChE Foundation and acknowledges funding from the Institute for Diabetes, Obesity, and Metabolism, the Penn Mental Health AIDS Research Center of the University of Pennsylvania, and the National Institute of General Medical Sciences of the National Institutes of Health under award number R35GM138201. IRB approval for this project was obtained from the University of Pennsylvania. Written informed consent was obtained from all participants and research/ethics approval and permits were obtained from the following institutions prior to sample collection: the University of Addis Ababa and the Federal Democratic Republic of Ethiopia Ministry of Science and Technology National Health Research Ethics Review Committee; COSTECH, NIMR and Muhimbili University of Health and Allied Sciences in Dar es Salaam, Tanzania; the University of Botswana and the Ministry of Health in Gaborone, Botswana; the Cameroonian National Ethical Committee (NEC) and Cameroonian Ministry of Health (MOH).

We acknowledge the Penn Medicine BioBank (PMBB) for providing data and thank the patient-participants of Penn Medicine who consented to participate in this research program. We would also like to thank the Penn Medicine BioBank team and Regeneron Genetics Center for providing genetic variant data for analysis. The PMBB is approved under IRB protocol# 813913 and supported by Perelman School of Medicine at University of Pennsylvania.

We thank Alex Harris, Alexander Platt, and Srilakshmi Raj for discussing the evolutionary analysis in the paper. We thank the following individuals and organizations for their essential work in collecting samples for this project: Kenya: Lilian A. Nyndodo, Eva Aluvalla, Daniel Kariuki, Fathya Abdo, and Hussein Musa; Ethiopia: Solomon Taye, Birhanu Mekaunintie, and Alemayehu Moges; Tanzania: Kweli Powell, Holly Mortensen, Mariki Euphrasia , Ruth Matiyas, John G. Memra, Holliness Santa, Emanuel Kimario, Reginald Kavishe; Botswana: Michael Campbells, Ari Ho-Foster, Maitseo M. M. Bolane, Maungo Moswang, Gaolape Mpoloka, Kingsley Motshegwe, Mothusi Molatlhegi; Cameroon: Eric Mbunwe, Sali Django, Dickson Ndizi, Valentine Ngum Ndze, Julius Fonsah, Eric Ngwang, Grace N. Tenjei, Meagan Rubel, Peter Kfu, BACUDA (Association Culturelle pour le Dévelopment Bagyeli/Bakola de l’Ocean, CADDAP (Centre d’Action pour le Dévelopment Durable des Autochtones Pygmées), MBOSCUDA (Mbororo Social and Cultural Development Association). We especially thank all African participants for their important contributions to this study.

## Regeneron Genetics Center Banner Author List and Contribution Statements

All authors/contributors are listed in alphabetical order.

## RGC Management and Leadership Team

Goncalo Abecasis, Ph.D., Aris Baras, M.D., Michael Cantor, M.D., Giovanni Coppola, M.D., Aris Economides, Ph.D., Luca A. Lotta, M.D., Ph.D., John D. Overton, Ph.D., Jeffrey G. Reid, Ph.D., Alan Shuldiner, M.D.

Contribution: All authors contributed to securing funding, study design and oversight. All authors reviewed the final version of the manuscript.

## Sequencing and Lab Operations

Christina Beechert, Caitlin Forsythe, M.S., Erin D. Fuller, Zhenhua Gu, M.S., Michael Lattari, Alexander Lopez, M.S., John D. Overton, Ph.D., Thomas D. Schleicher, M.S., Maria Sotiropoulos Padilla, M.S., Louis Widom, Sarah E. Wolf, M.S., Manasi Pradhan, M.S., Kia Manoochehri, Ricardo H. Ulloa.

Contribution: C.B., C.F., A.L., and J.D.O. performed and are responsible for sample genotyping. C.B, C.F., E.D.F., M.L., M.S.P., L.W., S.E.W., A.L., and J.D.O. performed and are responsible for exome sequencing. T.D.S., Z.G., A.L., and J.D.O. conceived and are responsible for laboratory automation. M.P., K.M., R.U., and J.D.O are responsible for sample tracking and the library information management system.

## Clinical Informatics

Nilanjana Banerjee, Ph.D., Michael Cantor, M.D. M.A., Dadong Li, Ph.D., Deepika Sharma, MHI.

Contribution: All authors contributed to the development and validation of clinical phenotypes used to identify study subjects and (when applicable) controls.

## Genome Informatics

Xiaodong Bai, Ph.D., Suganthi Balasubramanian, Ph.D., Andrew Blumenfeld, Boris Boutkov, Ph.D., Gisu Eom, Lukas Habegger, Ph.D., Alicia Hawes, B.S., Shareef Khalid, Olga Krasheninina, M.S., Rouel Lanche, Adam J. Mansfield, B.A., Evan K. Maxwell, Ph.D., Mrunali Nafde, Sean O’Keeffe, M.S., Max Orelus, Razvan Panea, Ph.D., Tommy Polanco, B.A., Ayesha Rasool, M.S., Jeffrey G. Reid, Ph.D., William Salerno, Ph.D., Jeffrey C. Staples, Ph.D.

Contribution: X.B., A.H., O.K., A.M., S.O., R.P., T.P., A.R., W.S. and J.G.R. performed and are responsible for the compute logistics, analysis and infrastructure needed to produce exome and genotype data. G.E., M.O., M.N. and J.G.R. provided compute infrastructure development and operational support. S.B., S.K., and J.G.R. provide variant and gene annotations and their functional interpretation of variants. E.M., J.S., R.L., B.B., A.B., L.H., J.G.R. conceived and are responsible for creating, developing, and deploying analysis platforms and computational methods for analyzing genomic data.

## Research Program Management

Marcus B. Jones, Ph.D., Michelle LeBlanc, Ph.D., Lyndon J. Mitnaul, Ph.D.

Contribution: All authors contributed to the management and coordination of all research activities, planning and execution. All authors contributed to the review process for the final version of the manuscript.

## Web Resources

Variation type descriptions in Variant Effect Predictor (VEP): https://uswest.ensembl.org/info/genome/variation/prediction/predicted_data.html

UCSC genome browser: https://genome.ucsc.edu/

