## Supplemental_Figures for "Impact of natural selection on global patterns of genetic variation, and association with clinical phenotypes, at genes involved in SARS-CoV-2 infection" (from another version of this preprint: version 1, 2021-07-01)

### Supplementary Figures

**Figure S1. Geographic information of ethnically diverse global populations included in our study.** Cameroon CAHG (Cameroon Pygmy): the Central African hunter-gatherers (CAHG) from Cameroon; EUR, European populations; EAS, East Asian populations; SAS, South Asian populations; AFR, African Niger-Congo populations; AMR, Native American populations; AA, African American populations; EA, European American populations. AA and EA are from the PMBB dataset, and EUR, EAS, SAS, AMR and AFR are from the 1000 genomes project. All other populations are from the TOPMed Africa6K project, and they are from five countries (Cameroon, Ethiopia, Kenya, Botswana, and Tanzania), belonging to four major different language families (Afroasiatic, Nilo-Saharan, Niger-Congo (or Niger-Kordofanian), and Khoesan). The Chabu (or Sabue) population from Ethiopia, the Hadza (or Hadzabe) population from Tanzania, the Sandawe population from Tanzania, and the Fulani population from Cameroon are listed as separate ancestral groups in our studies since their different evolutionary histories with other ethnic groups. The numbers in brackets denote sample sizes.

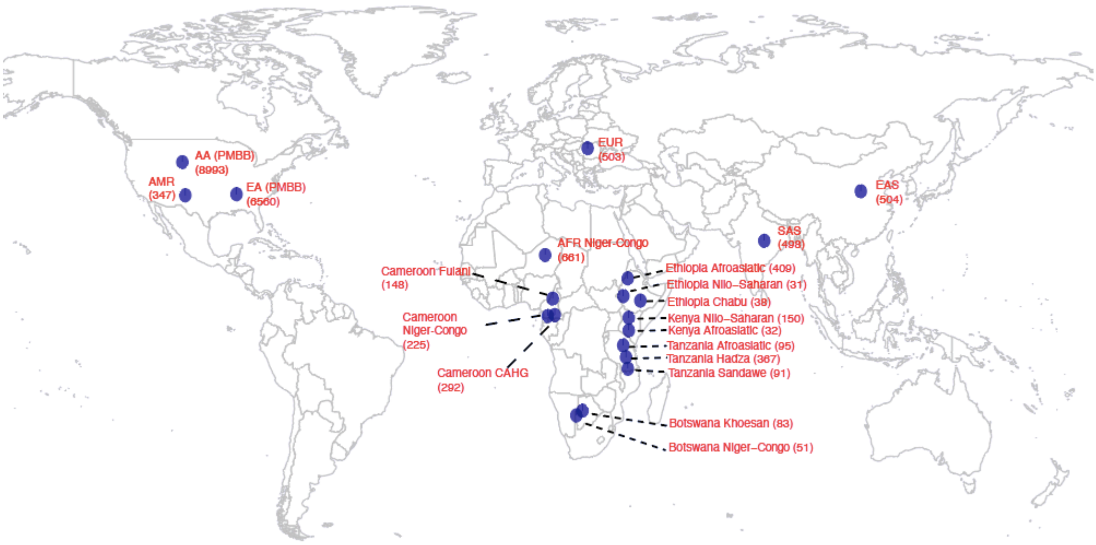

**Figure S2. MAF of six eQTLs identified at *ACE2*.**

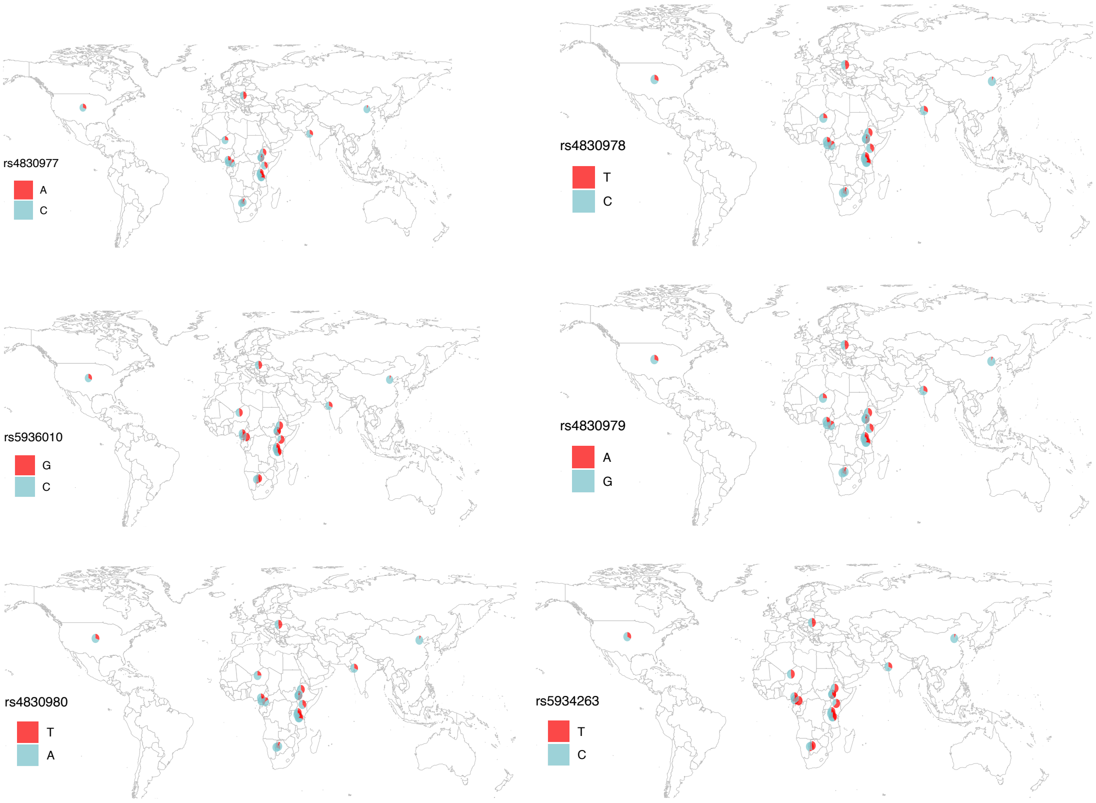

**Figure S3. Normalized expression data of the six eQTLs at *ACE2* in frontal cortex from the GTEx database.**

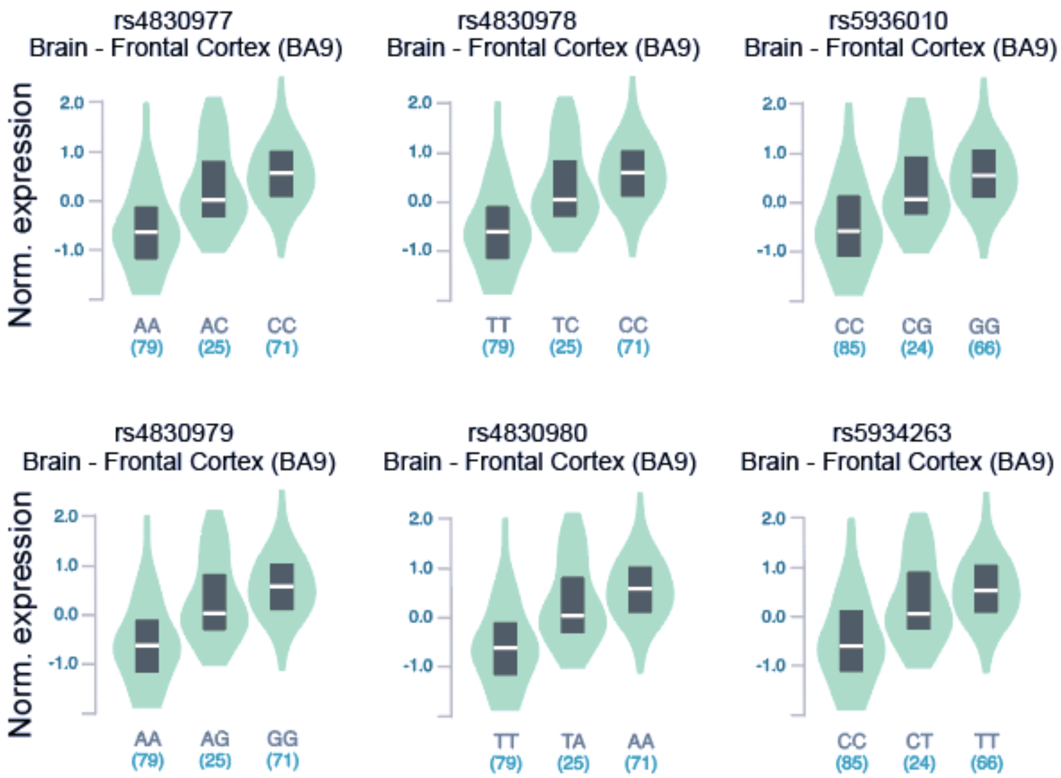

**Figure S4. Linkage disequilibrium between six eQTLs at *ACE2*.** $R^2$ were used to measure the LD.

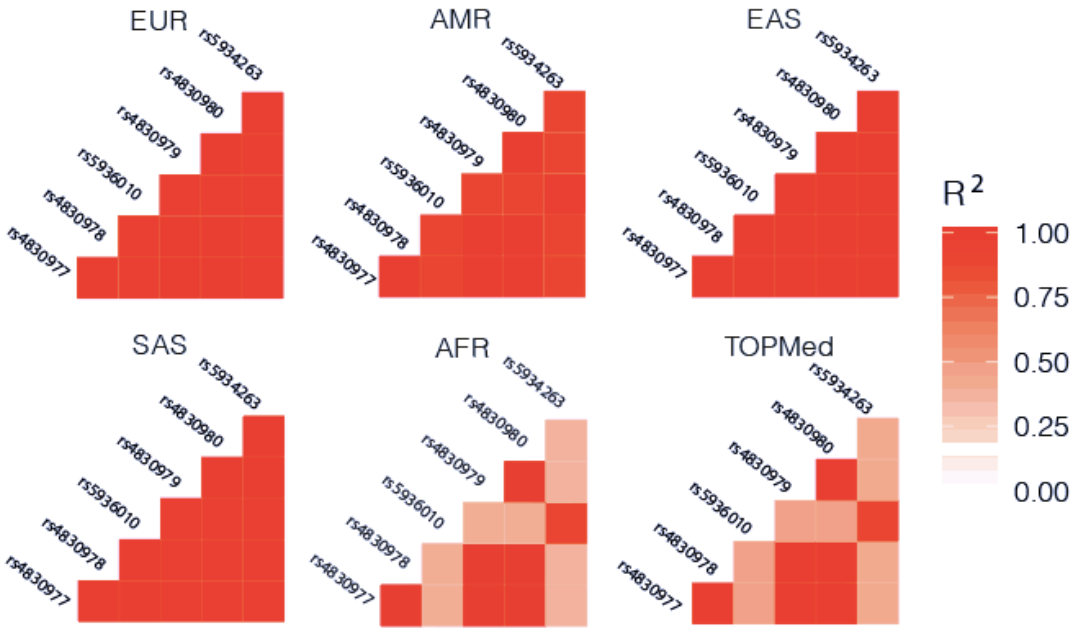

**Figure S5 Haplotype frequencies of the six eQTLs at *ACE2* in global populations.** Haplotypes with frequency <0.01 are not shown.

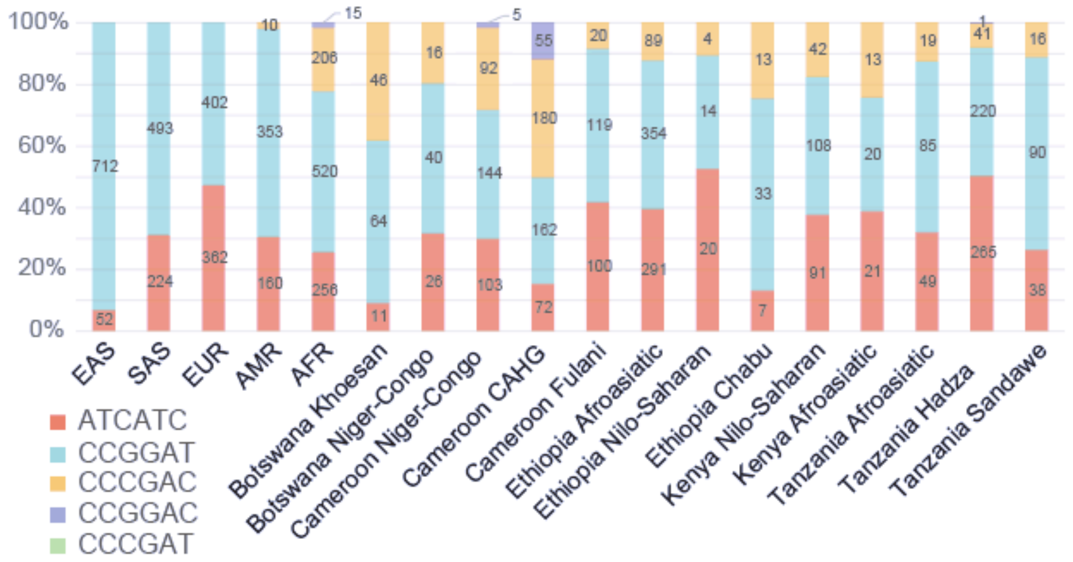

**Figure S6. Results of dN/dS test for the four candidate genes.** The dN/dS ratios of each ethnic group for *ACE2*, *TMPRSS2*, *DPP4*, and *LY6E* were plotted.

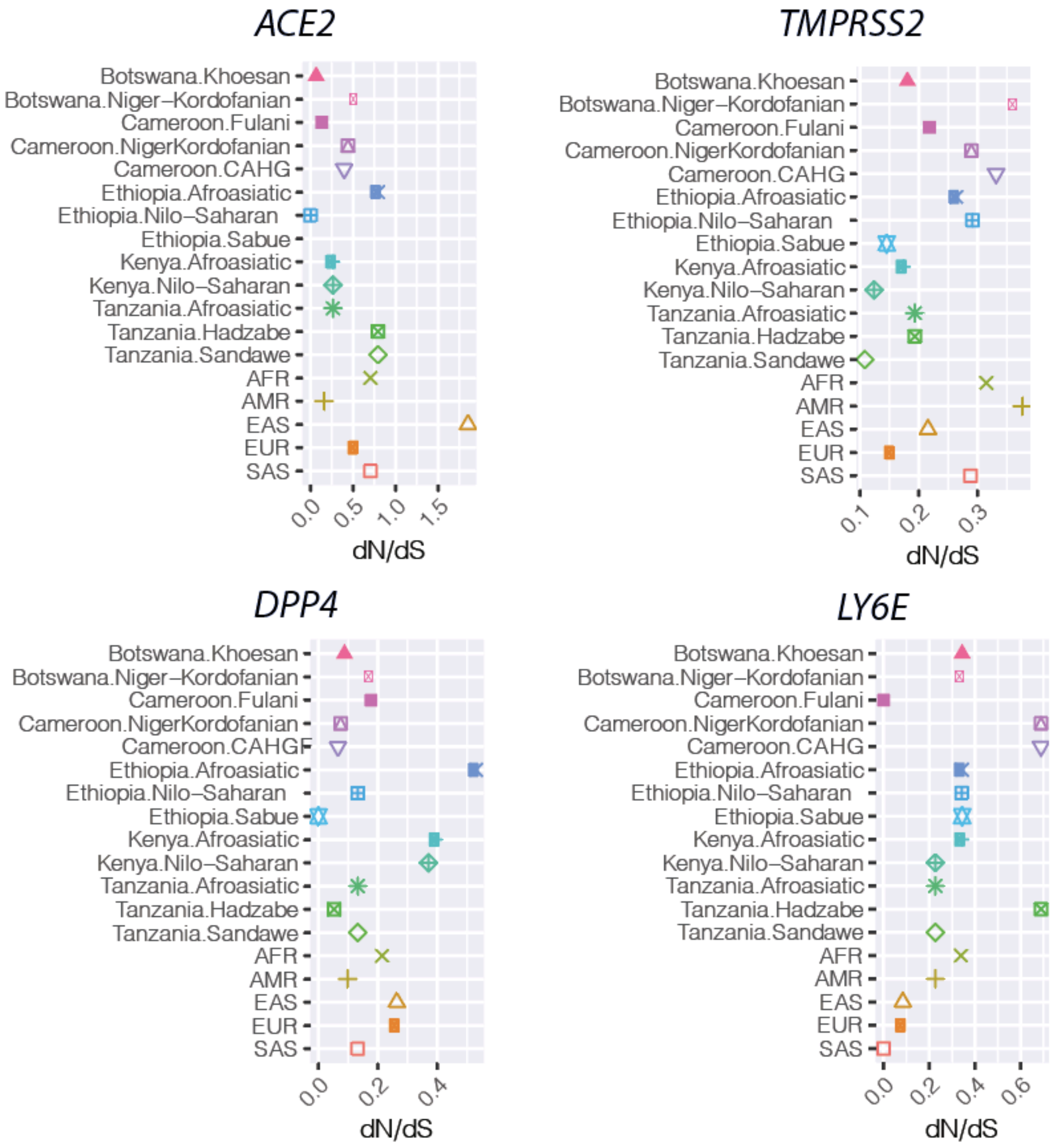

**Figure S7. Schematic illustration of the McDonald–Kreitman test.** Red dots denote non-synonymous variants and blue dots denote synonymous variants. Divergence of fixed variants mean that these variants were fixed in human lineage compared to the Chimpanzee. Polymorphism variants denotes that these variants were polymorphic within human populations. Dn, the number of divergence non-synonymous variants; Ds, the number of divergence synonymous variants; Pn, the number of polymorphism non-synonymous variants; Ps, the number of polymorphism synonymous variants; OR, odds ratio.

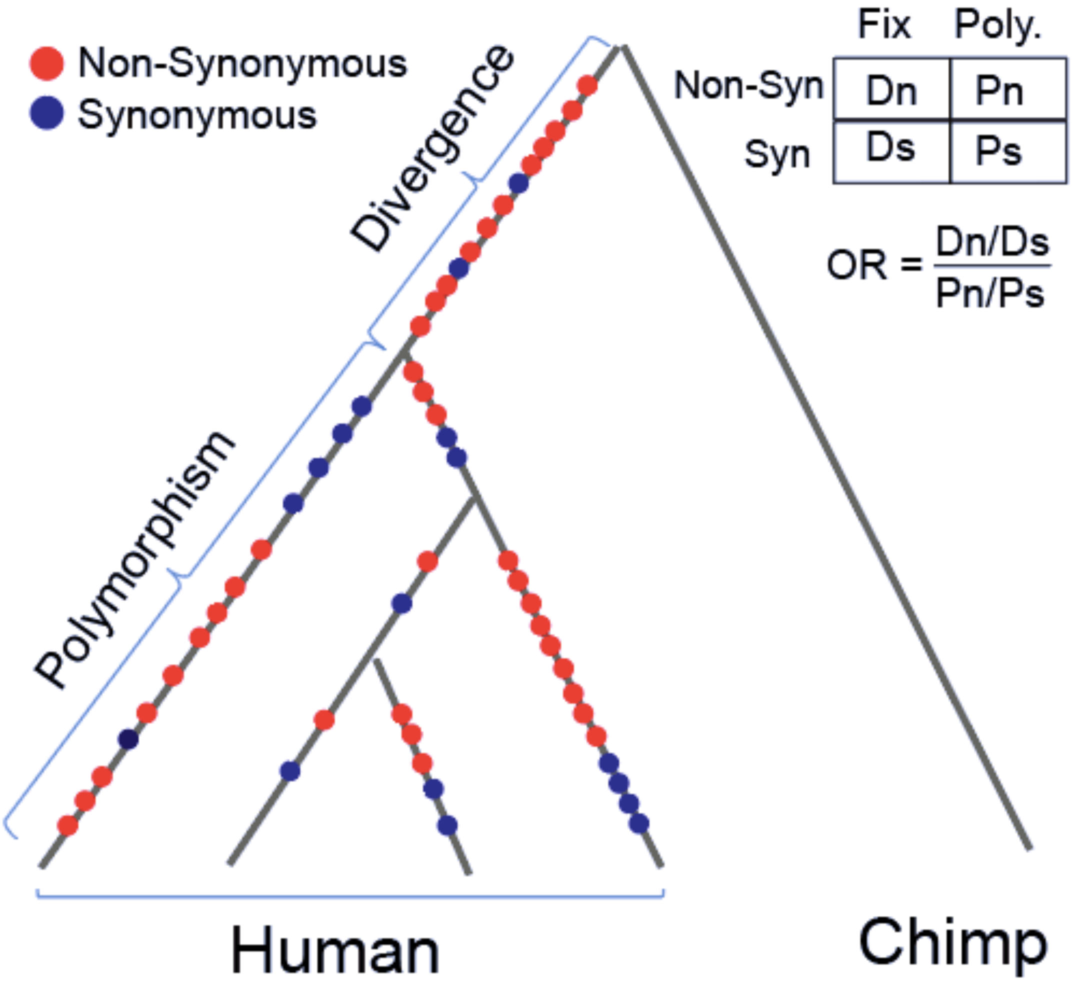

**Figure S8. Results of MK-test for four genes.** Odds ratios of (Dn/Ds) to (Pn/Ps) of each ethnic group for *ACE2* (A), *TMPRSS2* (B), *DPP4* (C), and *LY6E* (D) were plotted. Significance was tested by Fisher's exact test. No significant P-val was observed at three genes (*ACE2*, *DPP4* and *LY6E*). Odds ratios were not applied (NA) if no non-synonymous variants (Pn) were observed within individuals from a population.

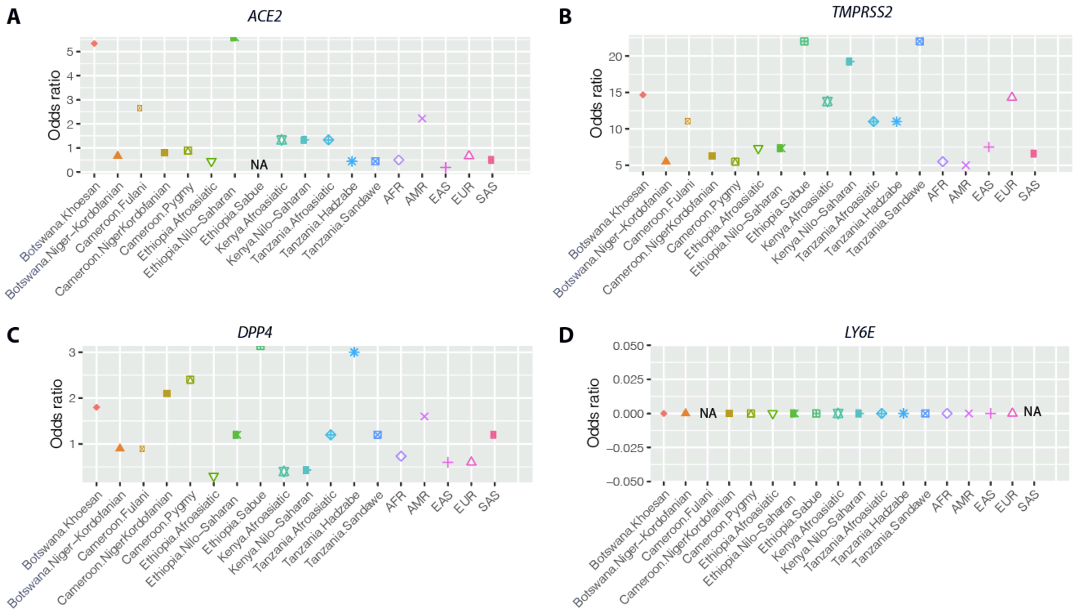

**Figure S9. The iHS scores for SNPs within *ACE2* region in each ethnic group.** Each dot represents a SNP. Dashed lines denote the empirical cutoff (|iHS|=2). Red dots mean that the corresponding SNPs harbor |iHS| scores > 2.

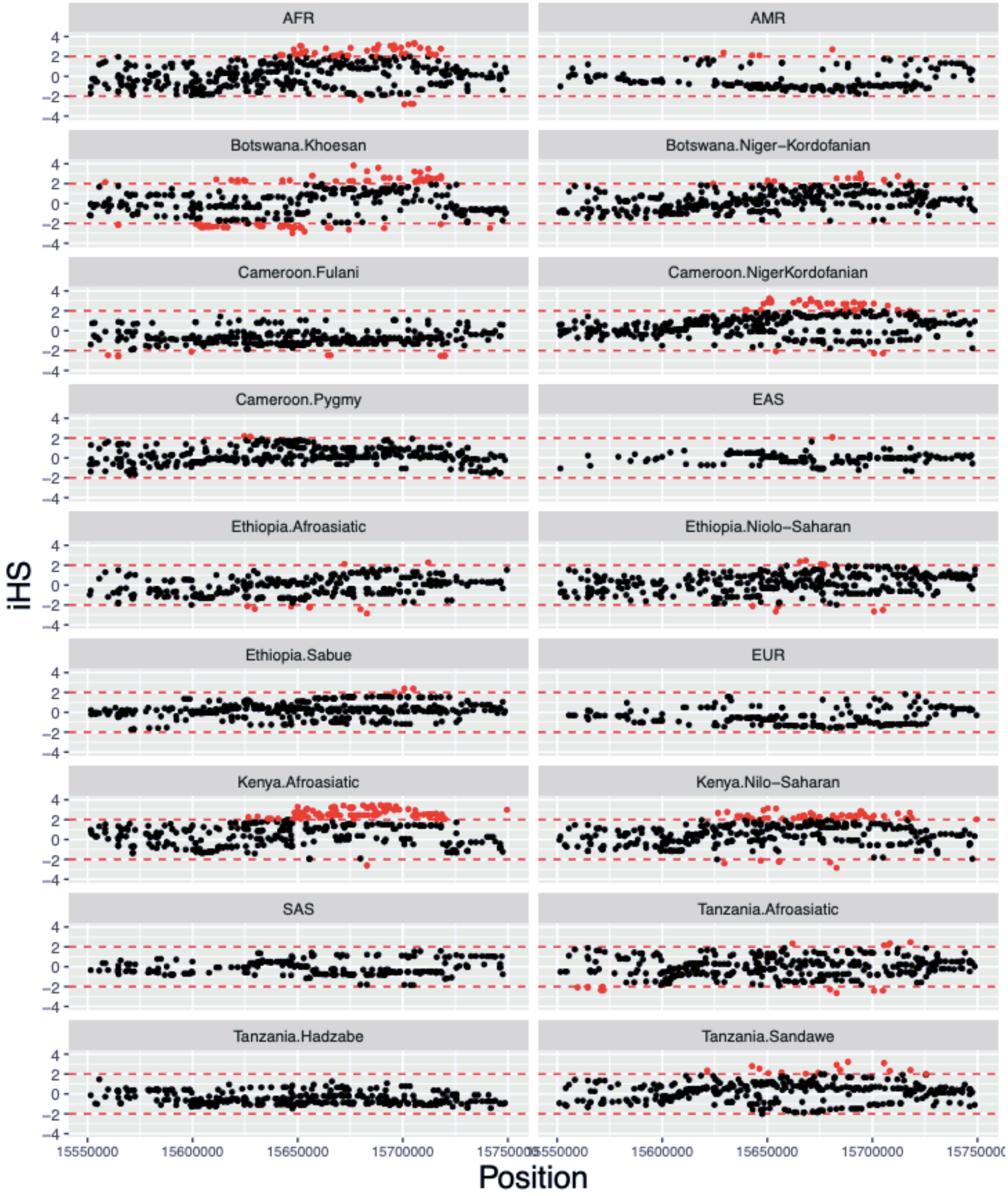

**Figure S10. LD pattern for all variants near *ACE2*.**

D prime was used to measure the LD. Dark gray tiles in the LD heatmap plot denote no variant was observed at the corresponding positions.

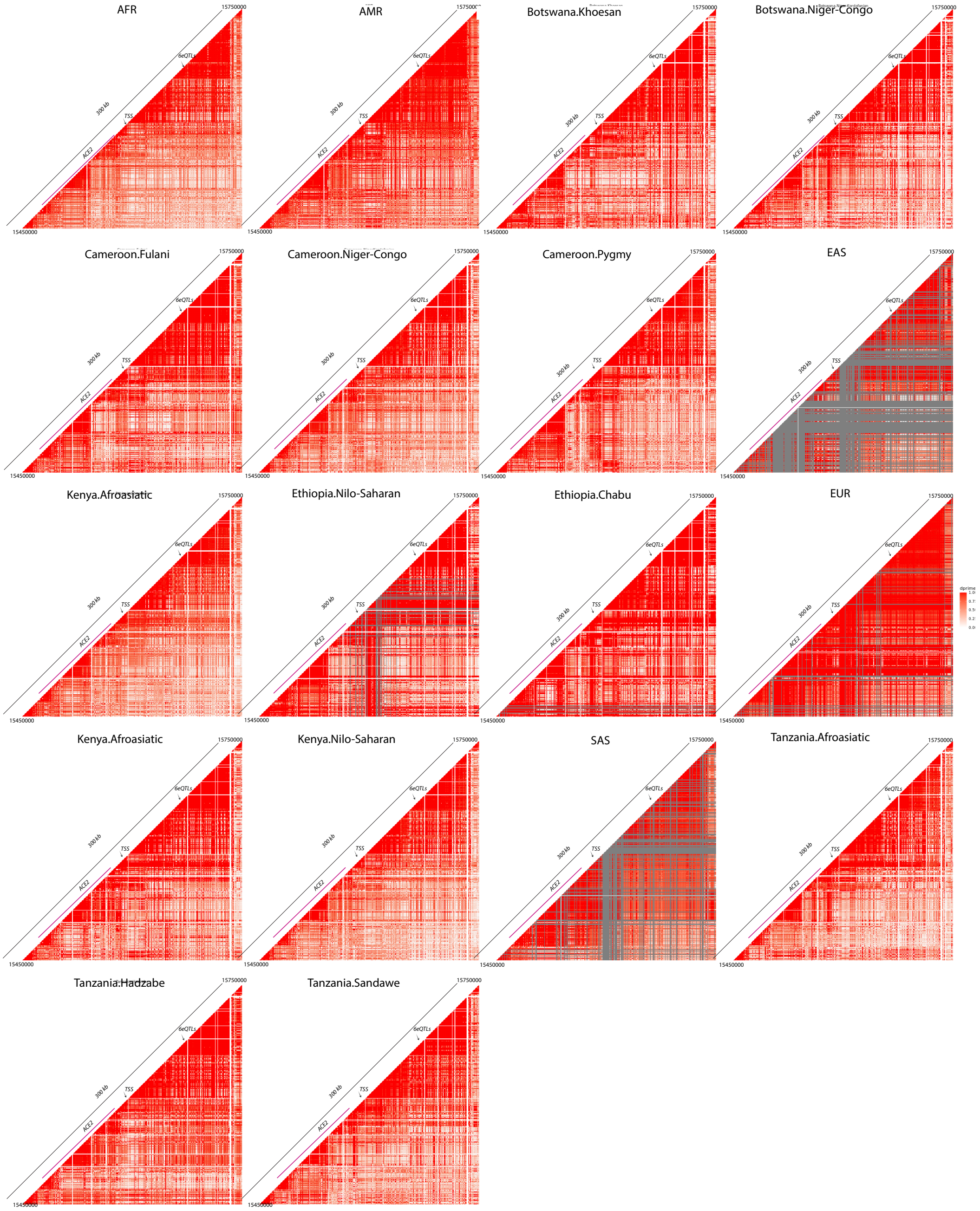

**Figure S11. LD pattern between selected variants near *ACE2*.** D prime was used to measure the LD. Variants included in the analysis are the four common variants (rs147311723, rs186029035, rs145437639, and rs138390800) identified in Cameroonian CAHG populations, 6 regulatory variants (rs4830977, rs4830978, rs5936010, rs4830979, rs4830980 and rs5934263), and four SNPs (rs150147953, rs2097723, rs4830984 and rs4830986) with significant selection signals at the upstream of *ACE2*. Dark gray tiles in the LD heatmap plot denote no variant was observed at the corresponding positions.

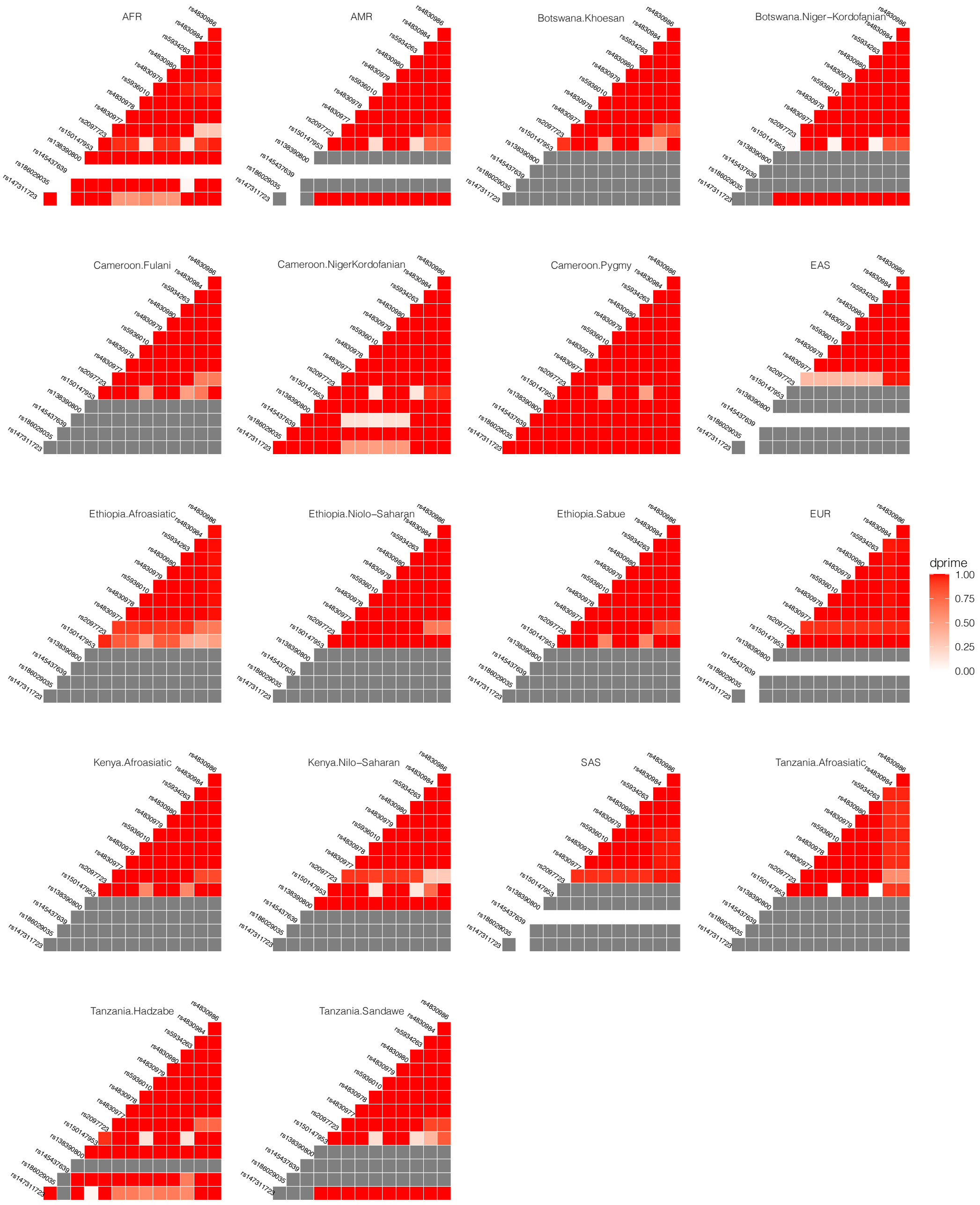

**Figure S12. The intersection of SNPs with significant selection signals and regulatory regions at *ACE2*.** SNPs with high iHS value (|iHS|>2) near *ACE2* locus overlapping DNase I hypersensitivity peaks from ENCODE (purple) or eQTLs from GTEx v8 (green) are shown in this figure. The SNPs discussed in the main text (rs150147953, rs2097723, rs5936010 and rs5934263) are highlighted with blue shadow. The DNase-seq tracks of large Intestine, small intestine, lung, kidney, heart, stomach, pancreas and skeletal muscle are also from ENCODE, and their signals are scaled to 1.5.

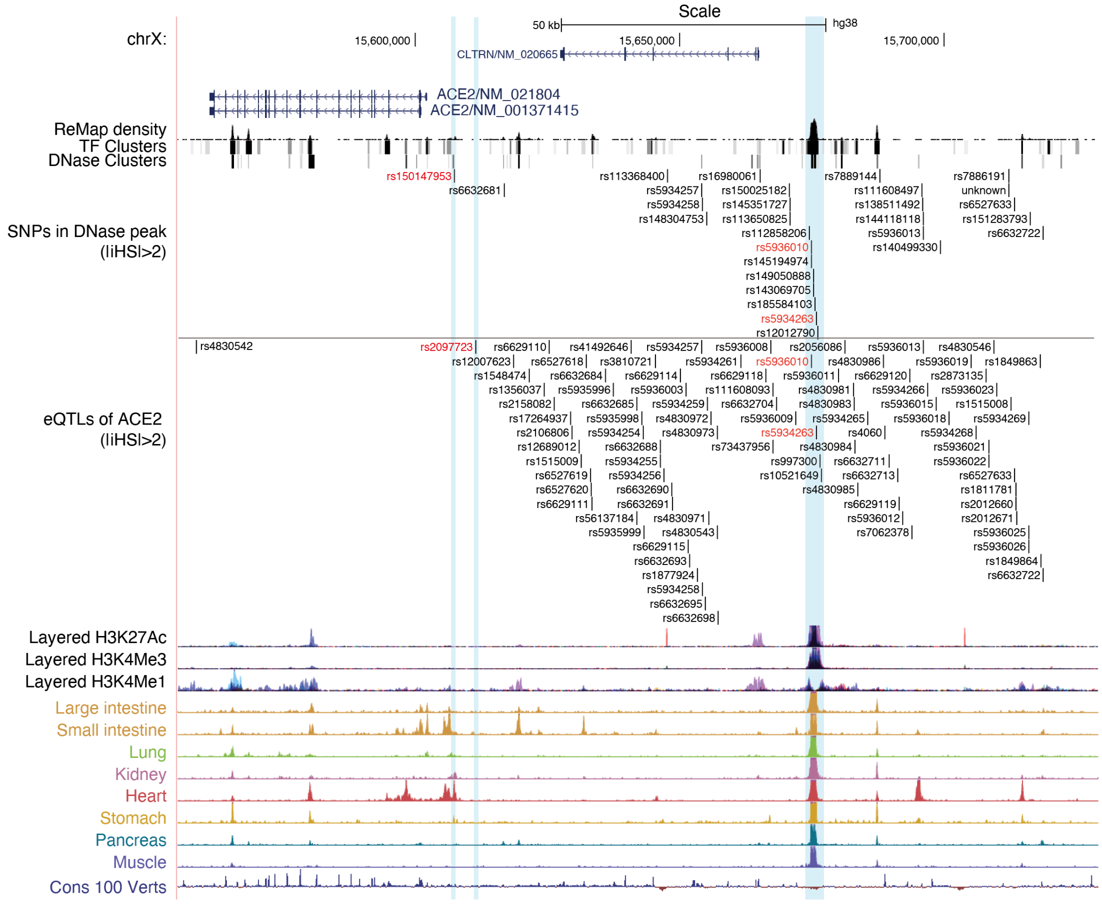

**Figure S13. Normalized expression data of the two eQTLs (rs76833541 and rs4283504) at *TMPRSS2* from the GTEx database.**

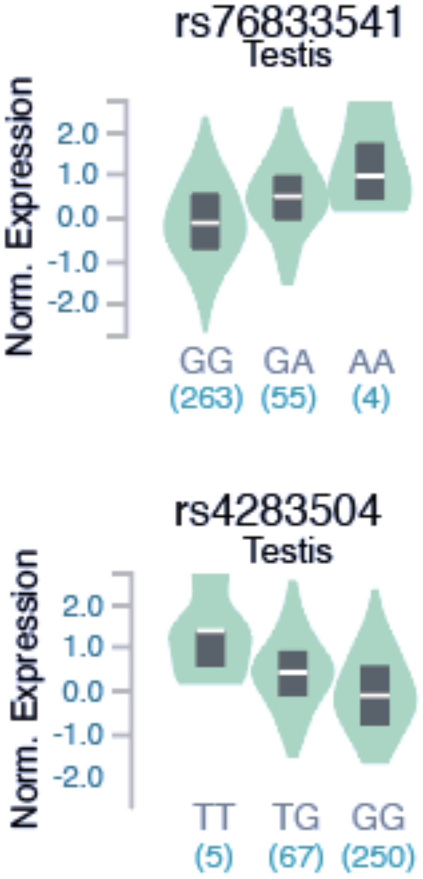

**Figure S14. MAF of two regulatory variants (rs76833541 and rs4283504) at *TMPRSS2*.**

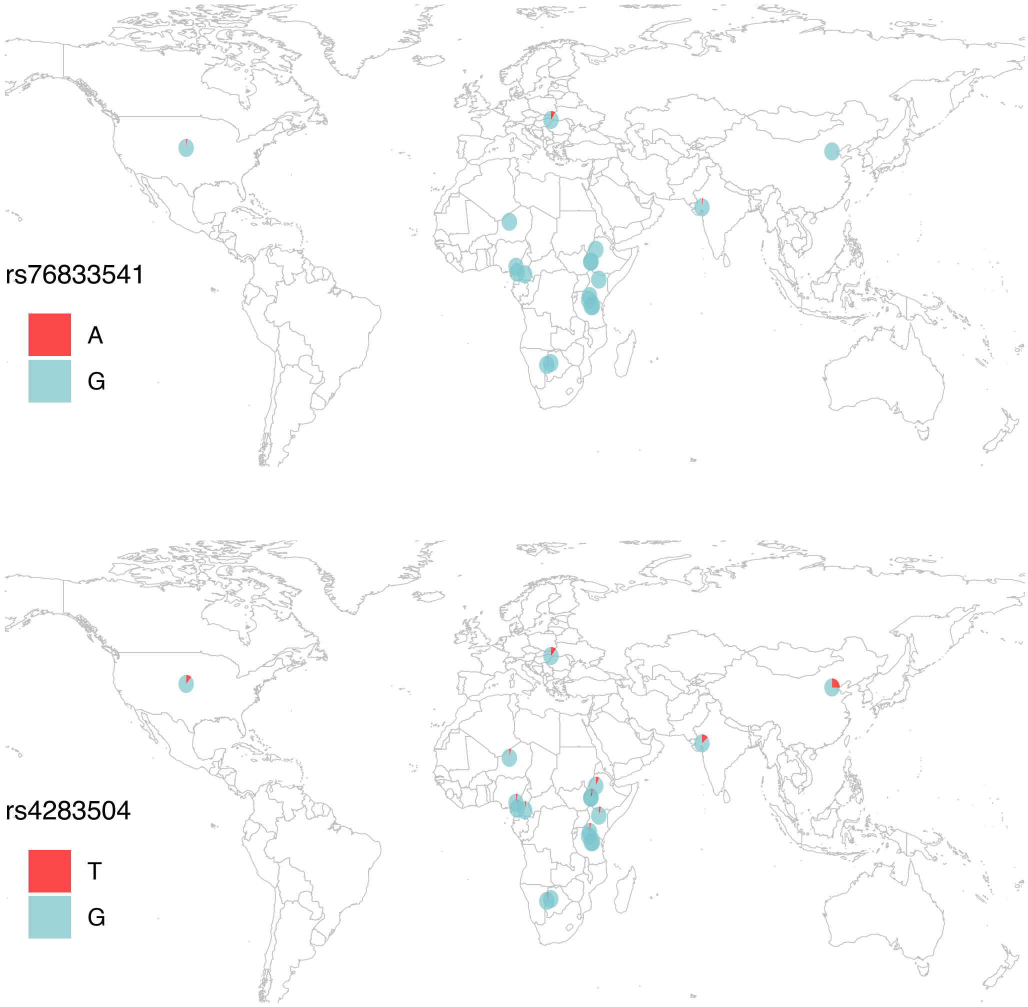

**Figure S15. MK-test for transcript ENST00000332149.10 of *TMPRSS2*.** There are 11 non-synonymous and 2 synonymous variants in ENST00000332149.10 that were fixed in human populations. These variants are located on different structure domain of *TMPRSS2*: amino acid T9P, A33V, R66C, and M67T in the cytoplasmic region; L87I in the transmembrane region; N107K in the extracellular region; S128N and S141G in the LDL-receptor class A domain; E374Q and T478M in the Peptidase S1 domain; S492G in the last residual); and A99A in the transmembrane region and G258G in the Peptidase S1 domain.

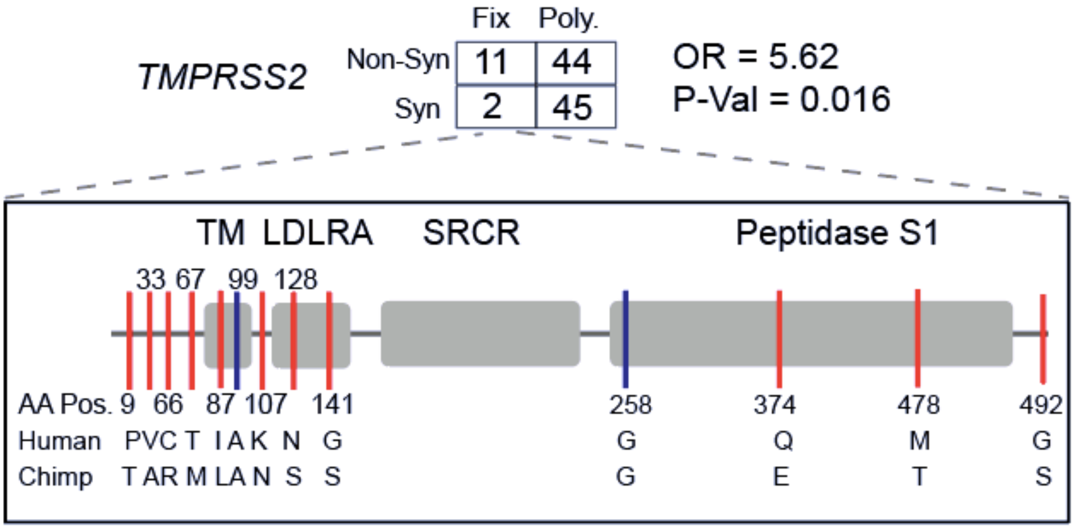

**Figure S16. iHS score for SNPs within *TMPRSS2* in each ethnic group.** Each dot represents a SNP. Dashed lines denote the empirical cutoff. Red dots mean that the corresponding SNPs harbor significant scores.

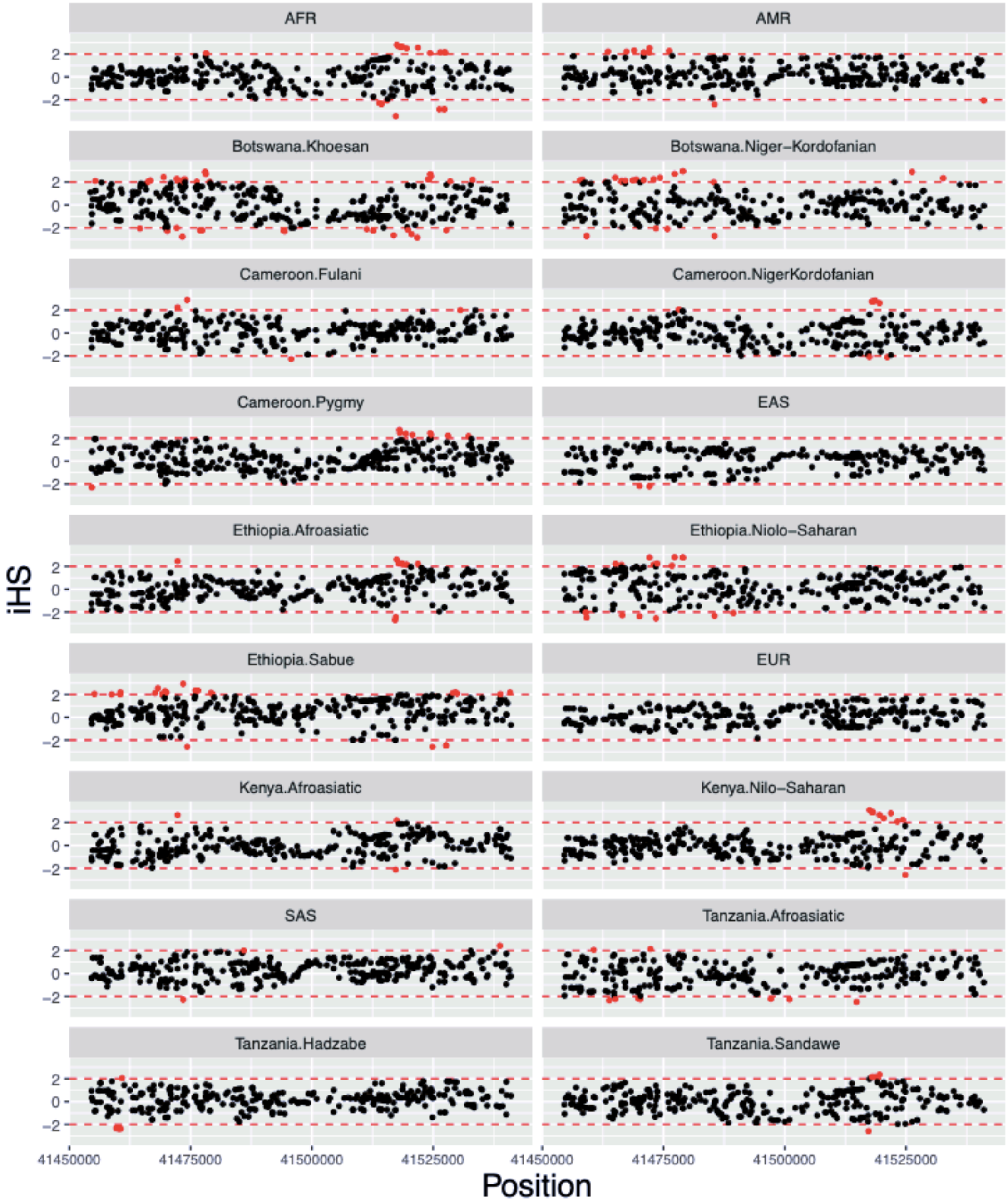

**Figure S17. LD pattern between 153 SNPs at *TMPRSS2* showing iHS signals in diverse ethnic groups.** D prime was used to measure the LD. Dark gray tiles in the LD heatmap plot denote no variant was observed at the corresponding position.

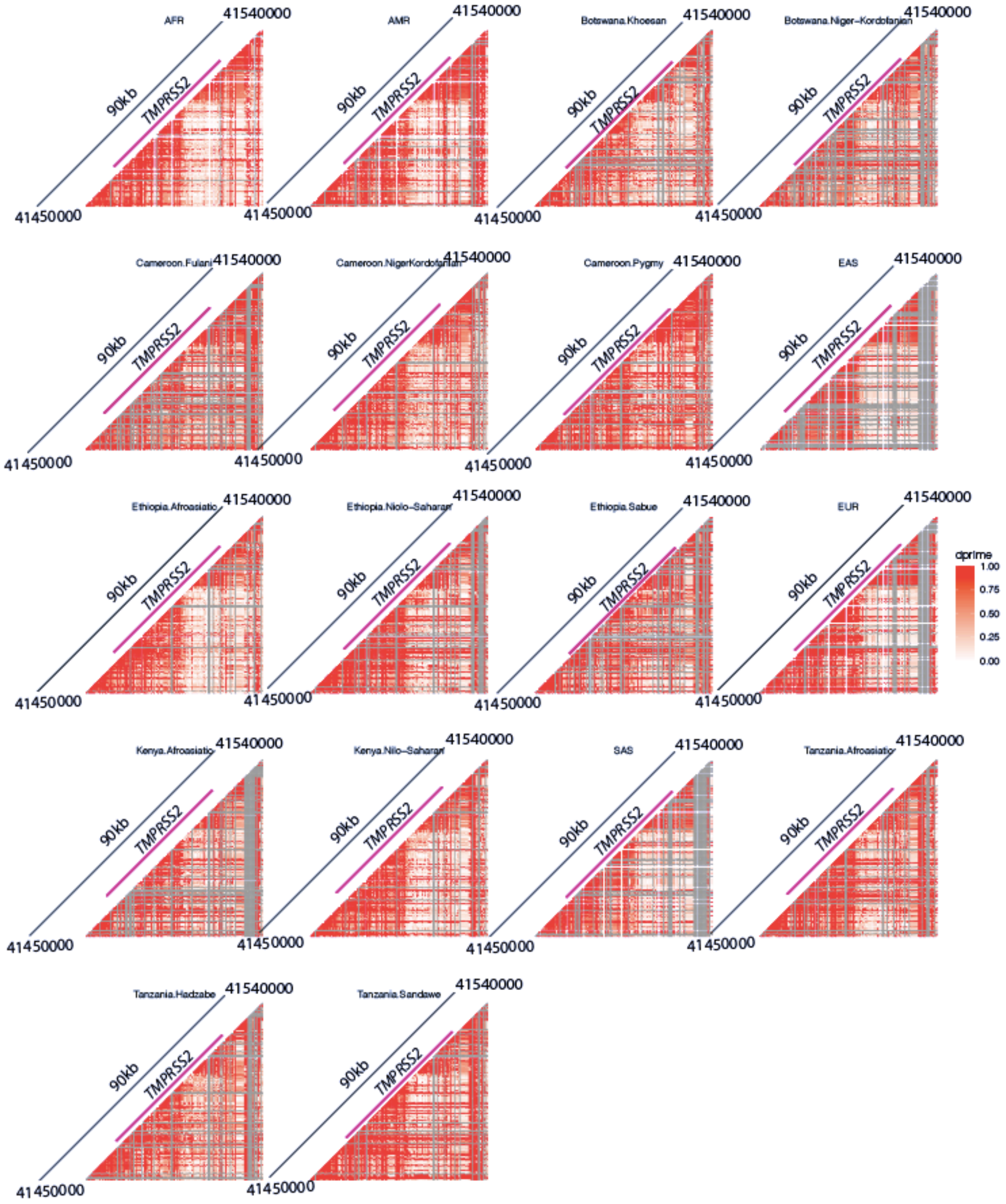

**Figure S18. LD pattern between selected variants (rs111870470, rs112306677, rs116170128, rs9636988, rs150969307 and rs73372191) at *TMPRSS2*.** D prime was used to measure the LD. Dark gray tiles in the LD heatmap plot denote no variant was observed at the corresponding position.

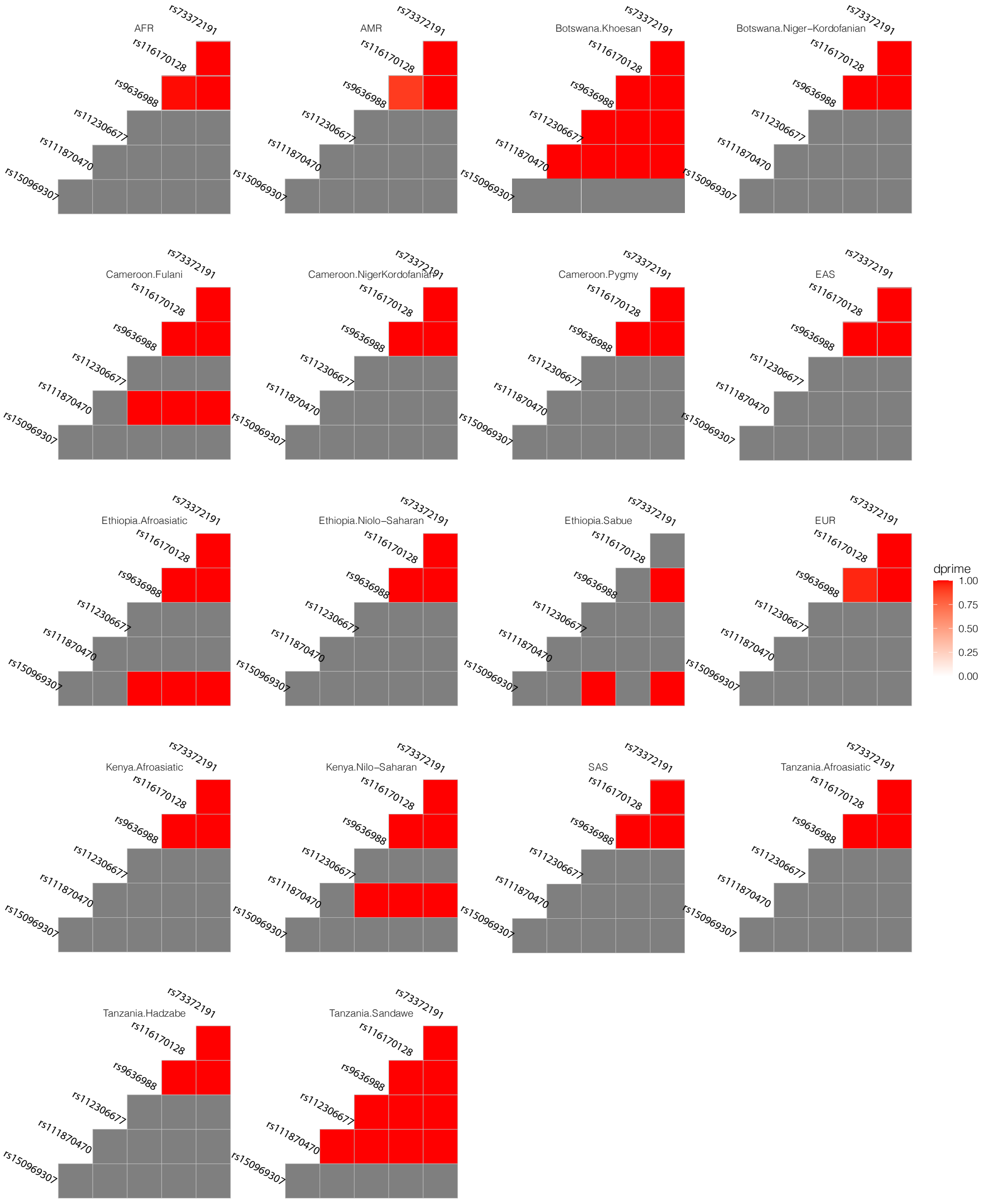

**Figure S19. The intersection of SNPs with significant selection signals and regulatory regions at *TMPRSS2*.** The SNPs rs435877, rs550390247, rs372713176, rs371744816, rs2838046 and rs777771526 are located in DNase peaks and transcription factor bind sites, and they are highlighted with light blue shadow. TF binding data are from ENCODE. Purple SNPs indicates the one with high iHS value (|iHS|>2) overlapping DNase I hypersensitivity peaks from ENCODE; green SNPs indicates they are significant eQTLs from GTEx v8 (green). The DNase-seq tracks of large Intestine, small intestine, lung, kidney, heart, stomach, pancreas and skeletal muscle are also from ENCOD, and their signals are scaled to 1.5.

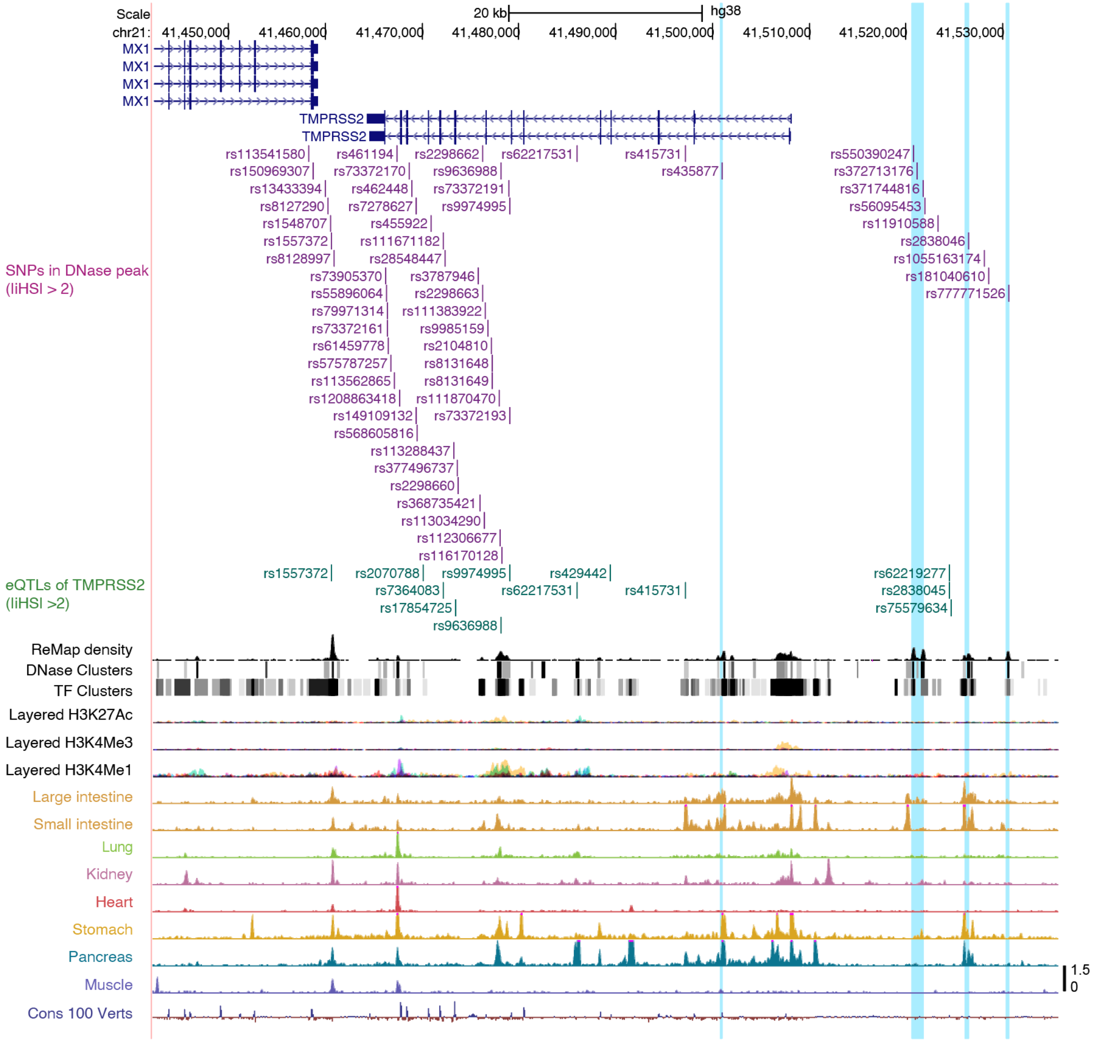

**Figure S20. Normalzied expression data from GTEx show the significant association between eQTL allele frequency and *DPP4* gene expression.**

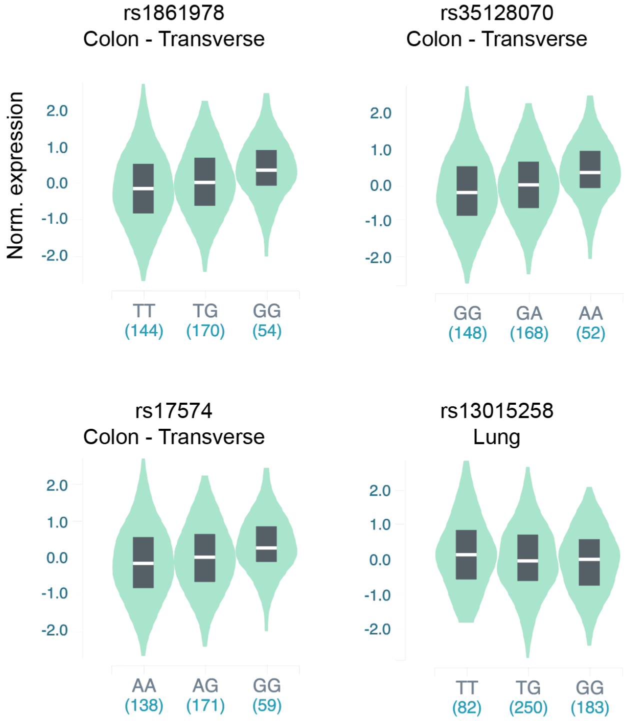

### Figure S21. MAF of four regulatory variants at *DPP4*

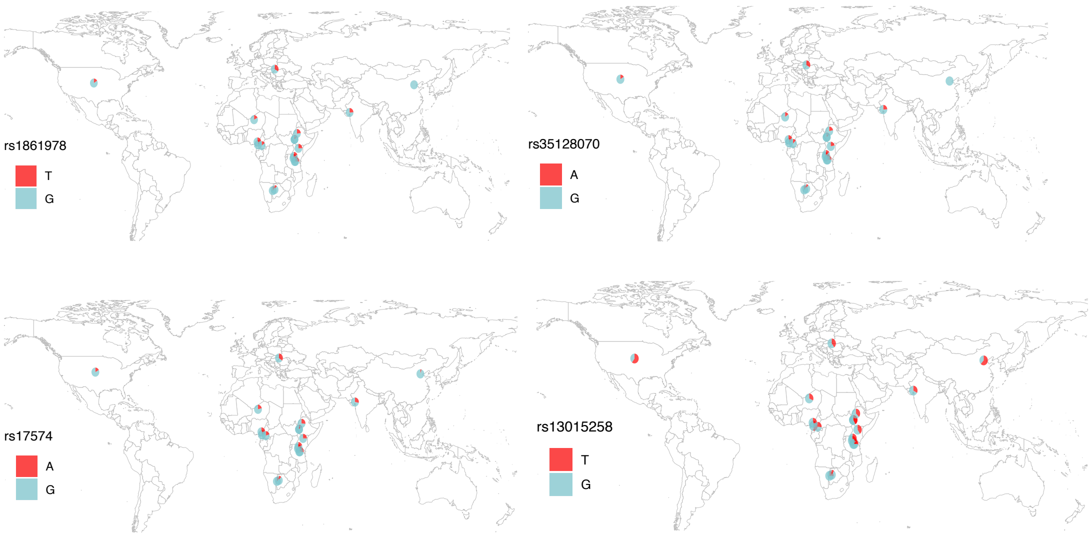

**Figure 22. iHS scores for SNPs at *DPP4*.** Each dot represents a SNP. Dashed lines denote the empirical cutoff. Red dots mean that the corresponding SNPs harbor significant scores.

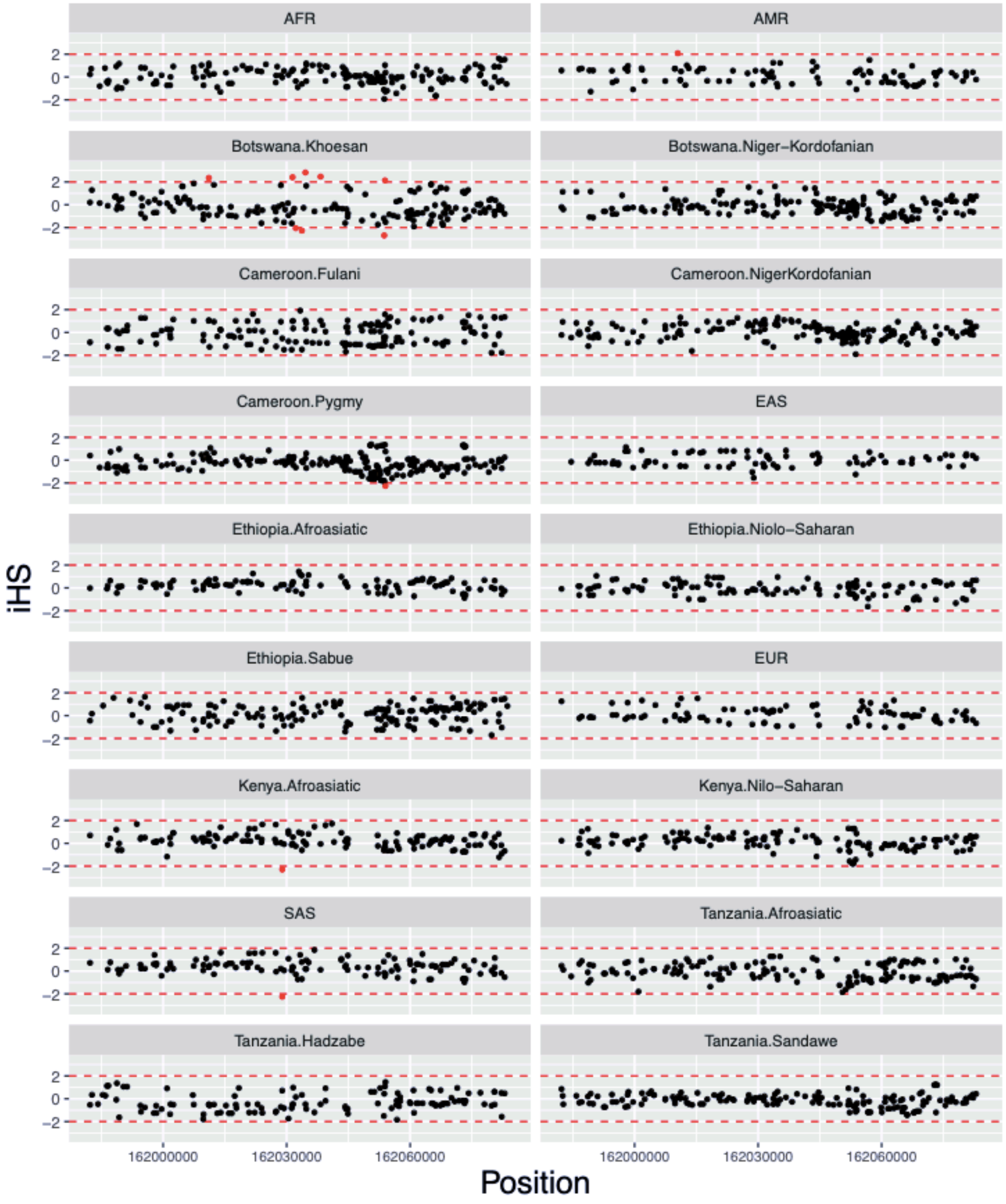

**Figure S23. LD pattern between 11 SNPs at *DDP4* showing iHS signals in diverse ethnic groups.** D prime was used to measure the LD. Eight of them were in the Khoesan populations from Botswana. Dark gray tiles in the LD heatmap plot denote no variant was observed at the corresponding positions.

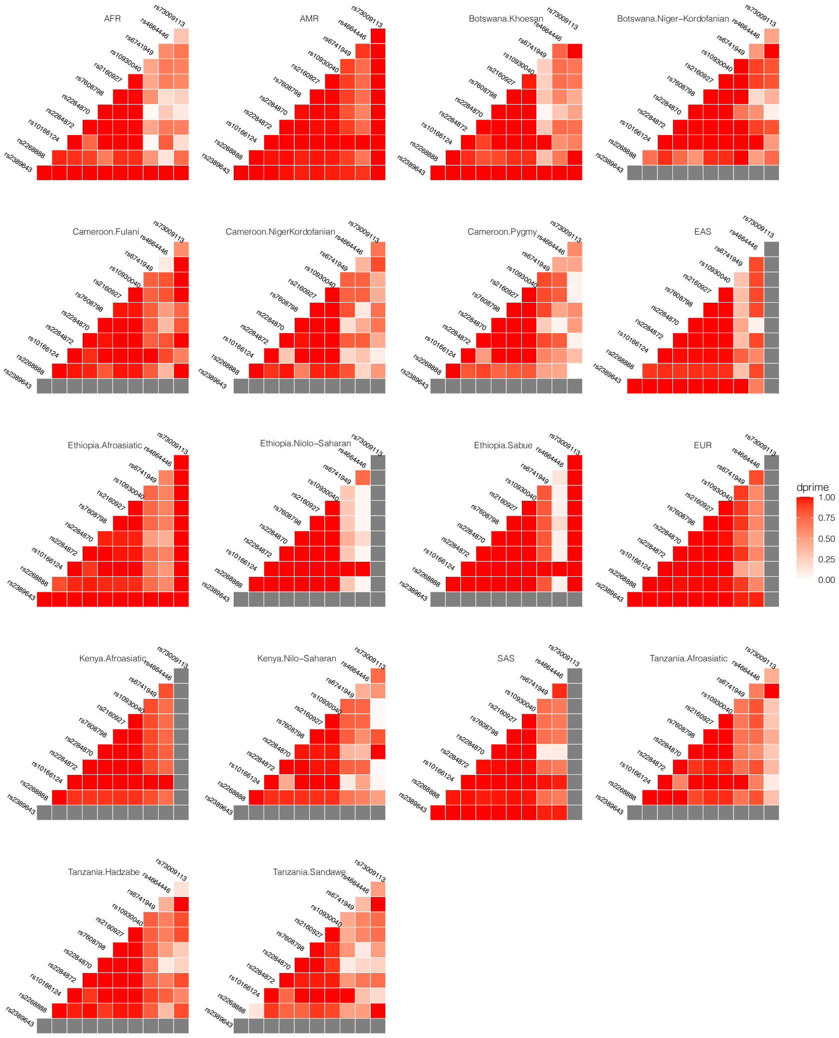

**Figure S24. The intersection of SNPs with significant selection signals and regulatory regions at *DPP4*.** The SNPs rs2098526 and rs2284870 highlighted with light blue shadow. Both SNPs have high iHS values and (|iHS|>2) overlap DNase I hypersensitivity peaks from ENCODE. The DNase-seq tracks of large Intestine, small intestine, lung, kidney, heart, stomach, pancreas and skeletal muscle are also from ENCODE, and their signals are scaled to 1.5.

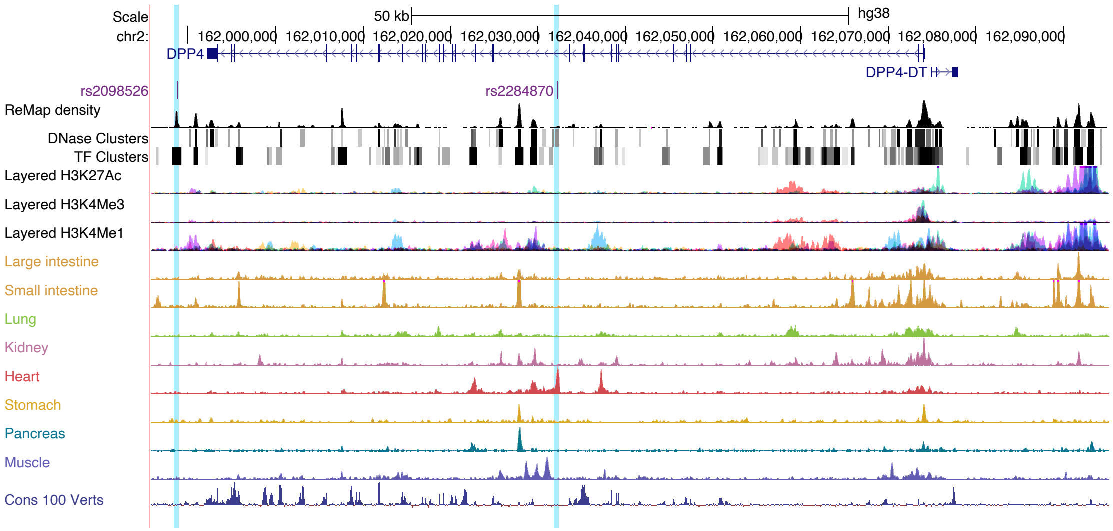

**Figure S25. Normalized expression data of the three eQTLs (rs13252884, rs17061979 and rs114909654) of *LY6E* in frontal cortex from GTEx database.**

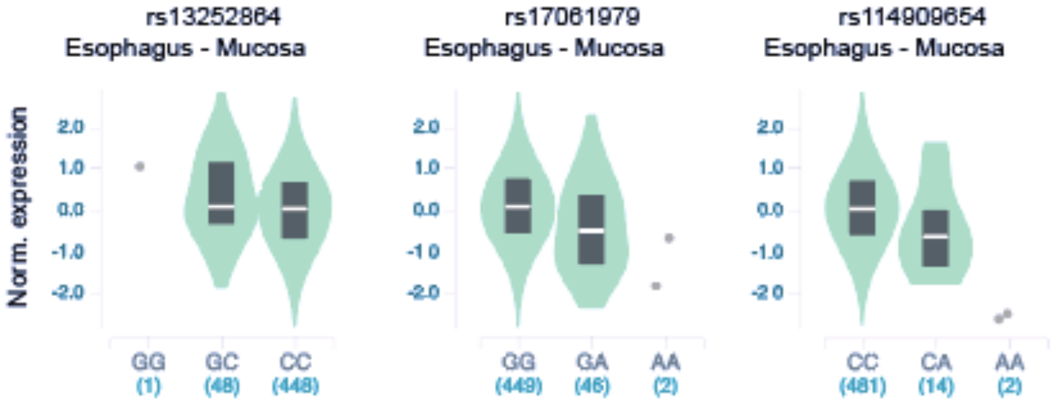

**Figure S26. MAF of three regulatory variants at *LY6E***

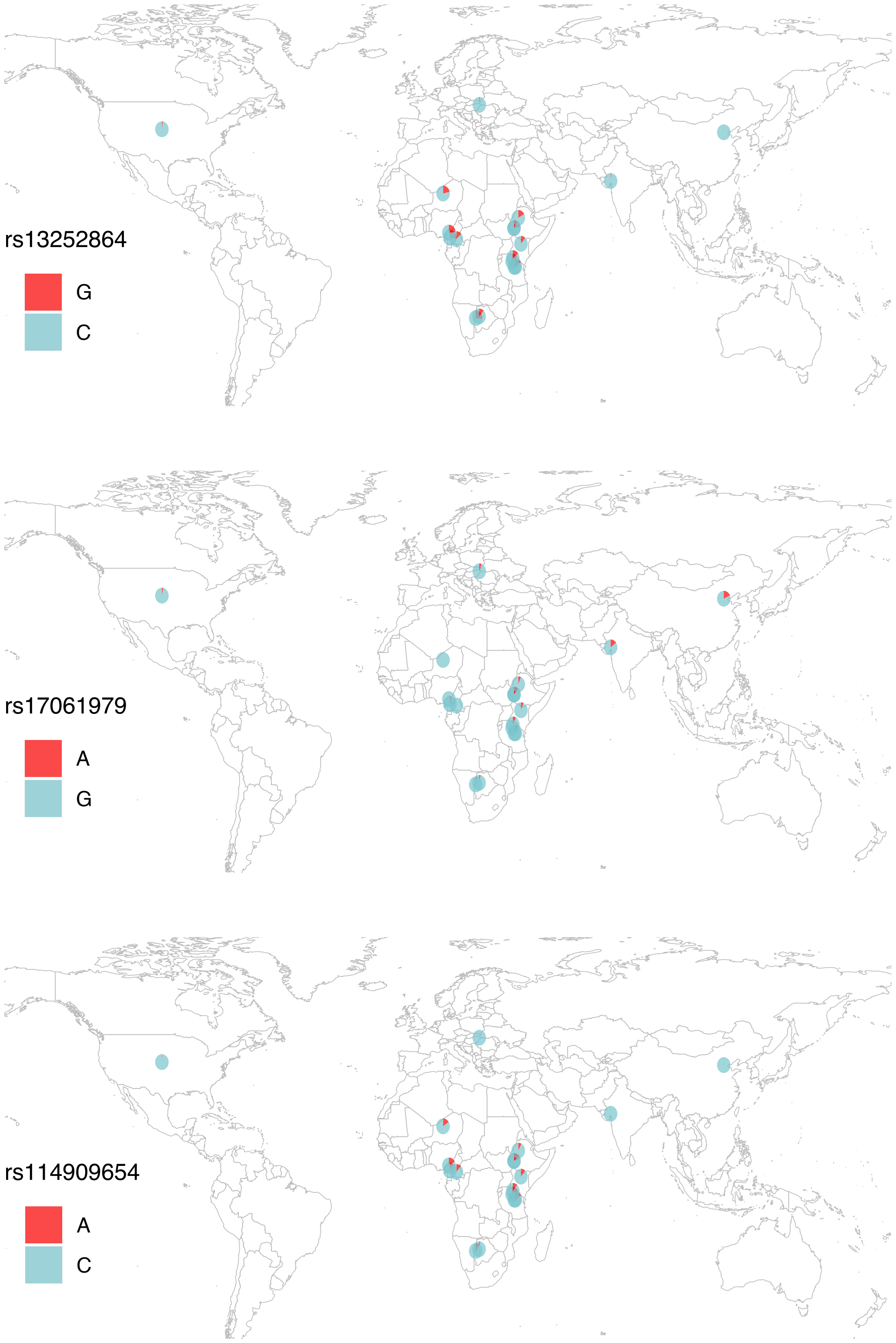

**Figure S27. iHS scores for SNPs at *LY6E*.** Each dot represents a SNP. Dashed lines denote the empirical cutoff. Red dots mean that the corresponding SNPs harbor significant scores.

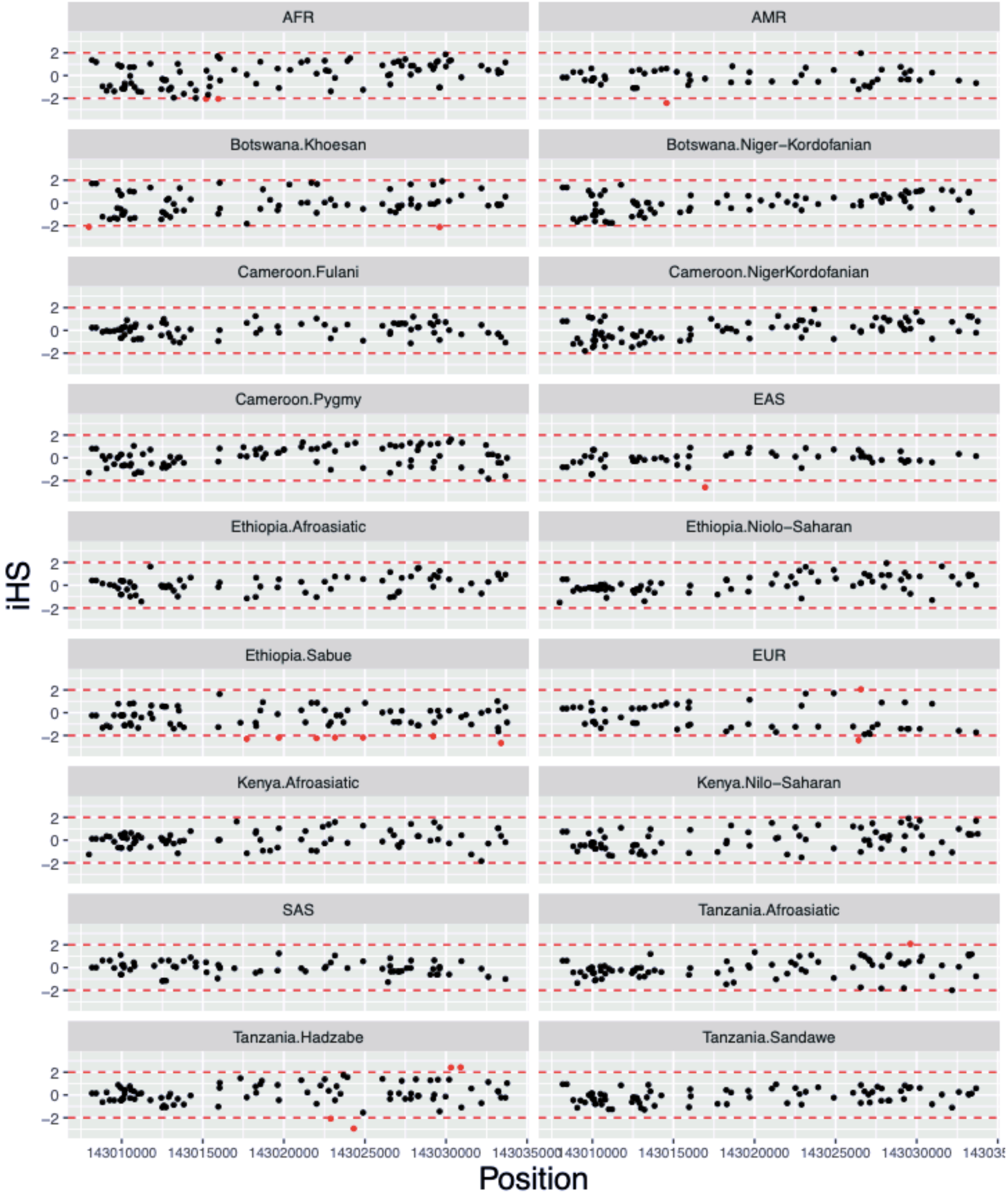

**Figure S28. LD pattern between SNPs at *LY6E* showing iHS signals in diverse ethnic groups.** D prime was used to measure the LD. Dark gray tiles in the LD heatmap plot denote no variant was observed at the corresponding positions.

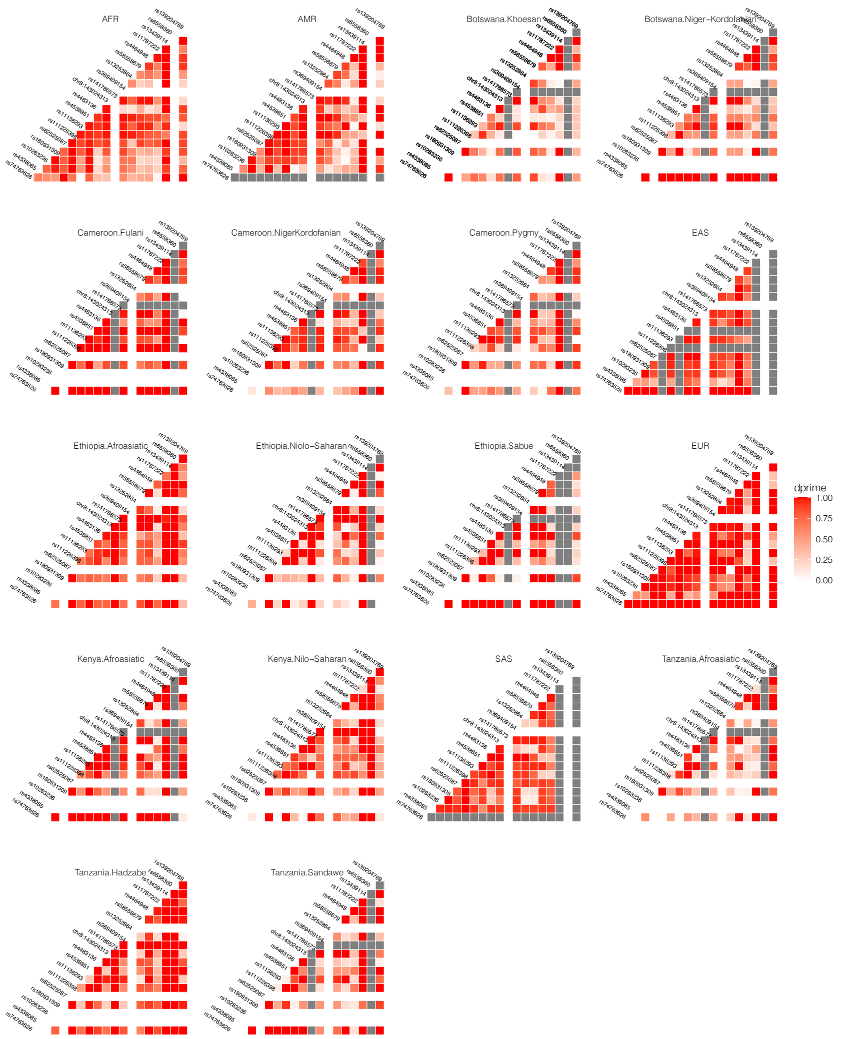

**Figure S29. The intersection of SNPs with significant selection signals and regulatory regions at *LY6E*.** SNPs with high iHS value (|iHS|>2) near DPP4 locus (< 10kb distance) overlapping DNase I hypersensitivity peaks from ENCODE (purple) or eQTLs from GTEx v8 (green) are shown in this figure. Potential regulatory elements are highlighted with blue shadow. The DNase-seq tracks of large Intestine, small intestine, lung, kidney, heart, stomach, pancreas and skeletal muscle are also from ENCODE.

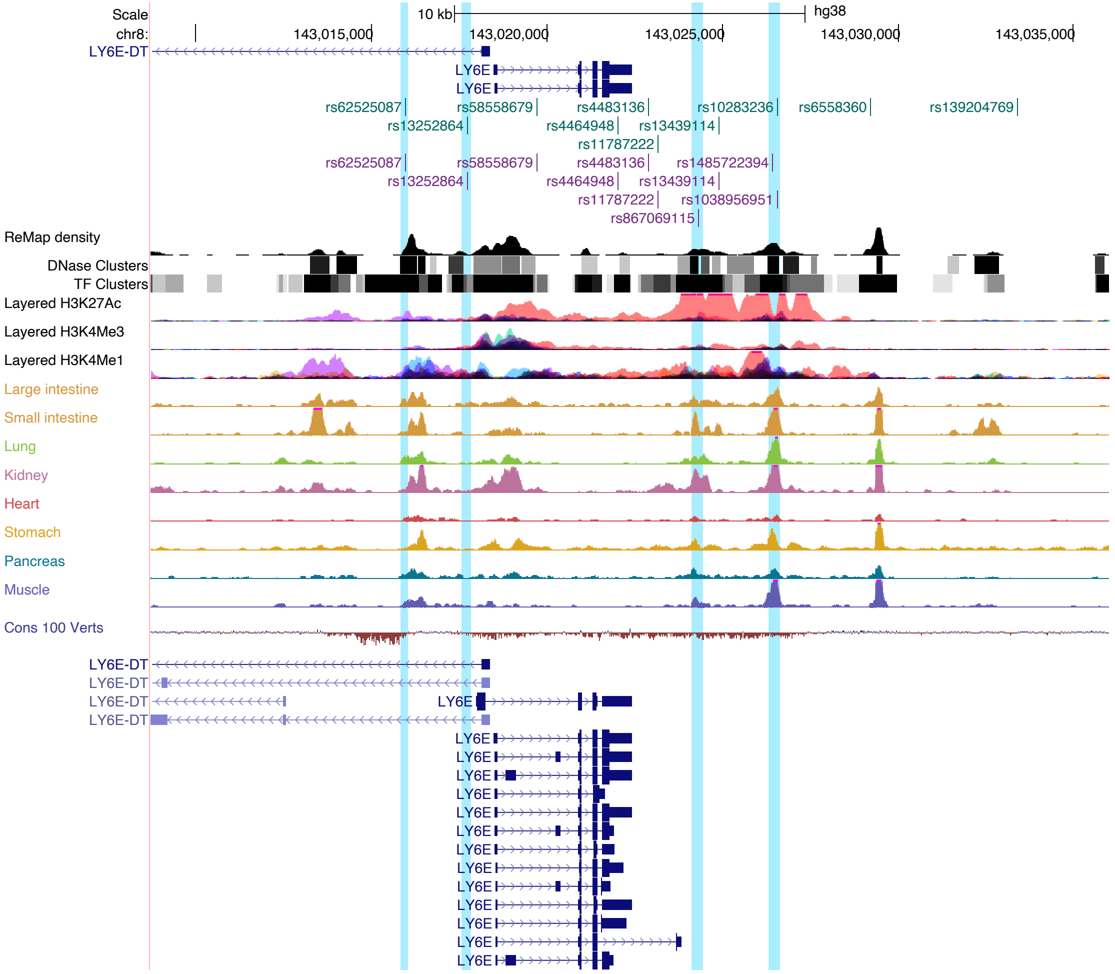
